# Disentangling shared genetic etiologies for kidney function and cardiovascular diseases

**DOI:** 10.1101/2024.07.26.24310191

**Authors:** Jun Qiao, Kaixin Yao, Yujuan Yuan, Xichen Yang, Le Zhou, Yinqi Long, Miaoran Chen, Wenjia Xie, Yixuan Yang, Yangpo Cao, Siim Pauklin, Jinguo Xu, Yining Yang, Yuliang Feng

## Abstract

Cardiovascular diseases (CVDs) are the leading cause of death worldwide, with chronic kidney disease (CKD) identified as a significant risk factor. CKD is primarily monitored through the estimated glomerular filtration rate (eGFR), calculated using the CKD-EPI equation. Although epidemiological and clinical studies have consistently demonstrated strong associations between eGFR and CVDs, the genetic underpinnings of this relationship remain elusive. Recent genome-wide association studies (GWAS) have highlighted the polygenic nature of these conditions and identified several risk loci correlating with their cross-phenotypes. Nonetheless, the extent and pattern of their pleiotropic effects have yet to be fully elucidated. We analyzed the most comprehensive GWAS summary statistics, involving around 7.5 million individuals, to investigate the shared genetic architectures and the underlying mechanisms between eGFR and CVDs, focusing on single nucleotide polymorphisms (SNPs), genes, biological pathways, and proteins exhibiting pleiotropic effects. Our study identified 508 distinct genomic locations associated with pleiotropic effects across multiple traits, involving 379 unique genes, notably *L3MBTL3* (6q23.1), *MMP24* (20q11.22), and *ABO* (9q34.2). Additionally, pathways such as stem cell population maintenance and the glutathione metabolism pathway were pivotal in mediating the relationships between these traits. From the perspective of vertical pleiotropy, our findings suggest a causal relationship between eGFR and conditions such as atrial fibrillation and venous thromboembolism. These insights significantly enhance our understanding of the genetic links between eGFR and CVDs, potentially guiding the development of novel therapeutic strategies and improving the clinical management of these conditions.

## Introduction

Cardiovascular diseases (CVDs) are the leading cause of mortality and disability globally, accounting for one-third of all deaths.^1^ The American Heart Association anticipates that by 2035, the economic impact of CVDs will escalate to approximately $1.1 trillion. Among the diverse risk factors, chronic kidney disease (CKD) is particularly significant, as it exacerbates heart disease.^2^ CKD leads to an increased production of reactive oxygen species (ROS) through oxidative stress, resulting in endothelial dysfunction—a critical precursor to atherosclerosis and an elevated risk of cardiovascular events.^3^ Affecting about 9.1% of the global population, as the World Health Organization reported in 2023, CKD is diagnosed and monitored by measuring the estimated glomerular filtration rate (eGFR), utilizing the CKD-EPI equation. An eGFR below 60 mL/min/1.73 m² signifies notable kidney impairment.^4,5^ Research by Go et al. has demonstrated a clear, graded association between declining eGFR and increased cardiovascular events.^6,7^ Additionally, studies focusing on older adults reveal that the risk of CVDs increases from 15% for those with an eGFR of 90 mL/min/1.73 m² to 40% for individuals at 30 mL/min/1.73 m² over three years.^8^ The Framingham Heart Study supports these findings, showing that individuals with mildly reduced eGFR levels (60 to 79 mL/min per 1.73 m²) face a higher risk of CVDs than those with higher eGFR.^9^ These studies underscore the significance of eGFR as a robust biomarker for assessing cardiovascular risk, highlighting its crucial role in clinical practice.

Genome-wide association studies (GWAS) have identified hundreds of variants contributing to the risk of eGFR and CVDs, some of which are shared risk loci. For example, Graham and colleagues pinpointed 147 loci associated with eGFR, seven demonstrating significant co-localization with cardiovascular traits, highlighting a genetic link between eGFR and various CVD phenotypes.^10^ Genetic pleiotropy, where a single genetic variant influences multiple traits, is prevalent in complex human diseases, particularly in loci associated with CVDs. This pleiotropy can be vertical, where a variant’s effect on one trait impacts another, or horizontal, where a variant independently affects several traits. Mendelian randomization (MR) analyses utilizing vertical pleiotropy have estimated causal relationships between eGFR and CVDs. For example, Kelly et al. established a causal link between declining eGFR and increased stroke risk,^11^ while Geurts et al. identified a bidirectional causal relationship between eGFR and atrial fibrillation (AF).^12,13^ However, recent studies suggest that eGFR does not significantly affect other CVDs, such as AF, coronary artery disease (CAD), stroke, and heart failure (HF). These inconsistencies in MR results, possibly due to residual biases and unmeasured confounders, necessitate further investigation. Furthermore, the study by Gong et al. highlights the role of horizontal pleiotropy in explaining the common genetic architecture of human phenotypes, within the context of recent advances in genomic statistical tools.^14^ Despite these advances, the extent of horizontal pleiotropy between eGFR and CVDs still needs to be explored, and the common genetic mechanisms must be fully elucidated. Therefore, there is a critical need for systematic research to investigate the shared genetic architectures between eGFR and CVDs, to identify shared polygenic risk variants, and to explore the involvement of specific molecular biological pathways.

In this study, we conducted an extensive analysis leveraged the latest GWAS summary statistics for European ancestry to explore the genetic associations between eGFR and six major CVDs, including AF, CAD, Venous Thromboembolism (VTE), HF, Peripheral Artery Disease (PAD), and Stroke. Firstly, the shared genetic basis is determined by quantifying the overall and local genetic correlations and exploring the overall genetic overlap. On this common genetic basis, in-depth research was conducted on the potential genetic mechanisms underlying these traits. Regarding horizontal pleiotropy, first, pleiotropy variation at the single nucleotide polymorphism (SNP) level is detected, and then co-localization analysis is performed to locate candidate causal variations accurately. Subsequently, position mapping and expression quantitative trait loci (eQTL) mapping were used at the gene level to identify candidate multi-effect genes. In addition, we conducted enrichment analysis on biological pathways and elucidated the biological functions of genes associated with multiple effector sites. We also investigated the potential association between plasma protein levels and disease susceptibility and evaluated its potential as a therapeutic target.

Finally, MR analysis was conducted at the level of vertical pleiotropy, using whole genome SNP data to estimate bidirectional causal effects while considering confounding factors and sample overlap.In conclusion, our comprehensive analysis of pleiotropy revealed the intricate genetic interactions between eGFR and CVDs. This work provides a promising path for a deeper understanding of their genetic associations, laying a solid foundation for future studies and driving new insights and targets in prevention, early diagnosis, and personalized treatment.

## Methods

### Study Design

Figure 1 presents the workflow for this study.

**Figure 1:**
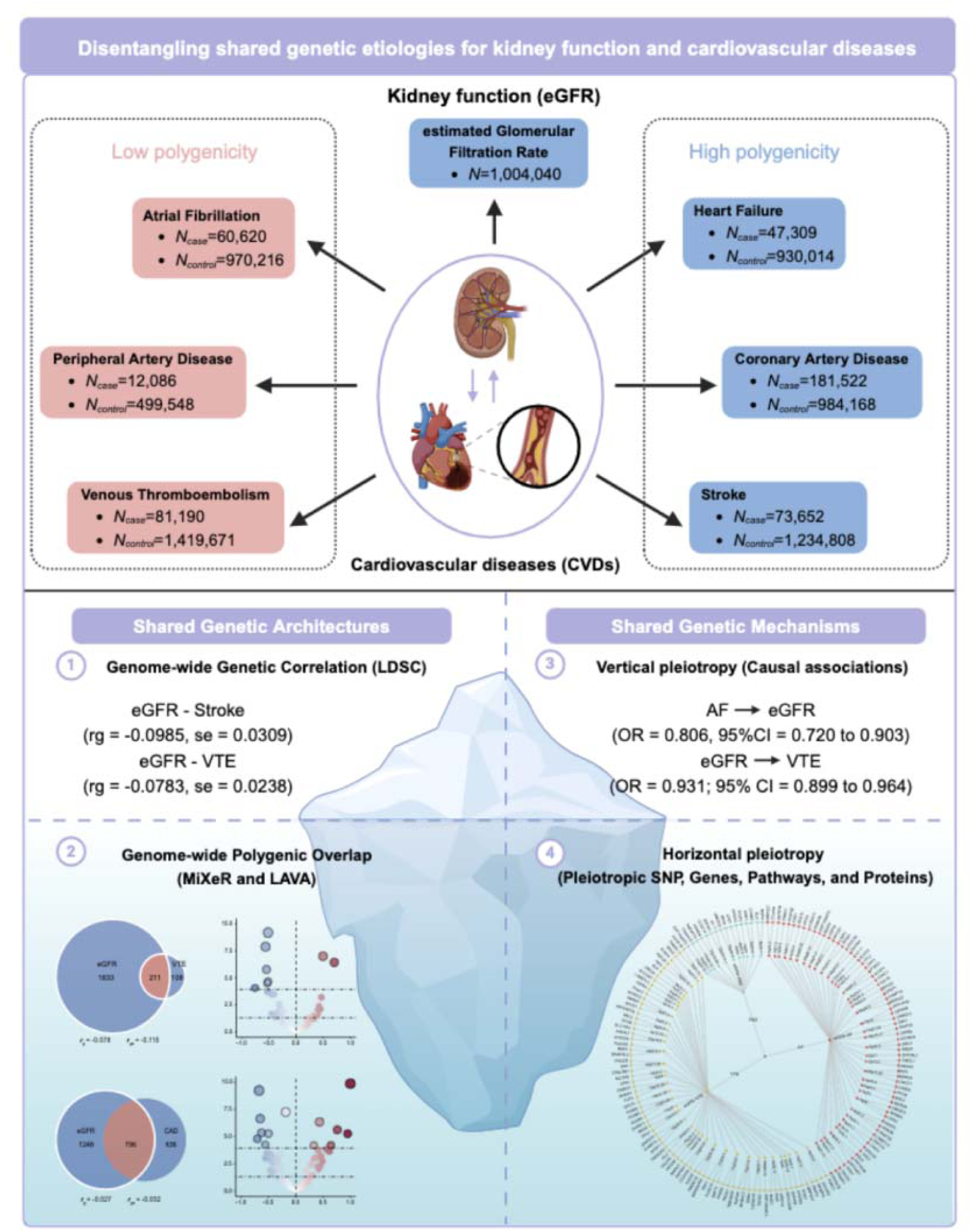
Schematic diagram of the analysis of genetic associations between glomerular filtration rate and six major cardiovascular diseases in this study. We analyzed the most comprehensive GWAS summary statistics to investigate the shared genetic architecture and potential mechanisms between eGFR and CVDs. First, the shared genetic basis was determined by quantifying global and local genetic correlations and exploring the global genetic overlap. Then, their two genetic pleiotropy were analyzed to investigate the shared genetic architecture, which can be vertical pleiotropy, that is, the effect of the variant on one trait affects another trait or horizontal pleiotropy, that is, the variant affects multiple traits independently. We first used Mendelian randomization on vertical pleiotropy to illustrate their causal relationship. Then, various statistical genetic methods were used on horizontal pleiotropy to sequentially apply pleiotropy analysis at the SNP, gene level, biological pathway, and protein target level to investigate the common genetic mechanism. Through this comprehensive genetic pleiotropy analysis, our understanding of the genetic link between eGFR and CVDs has been dramatically enhanced, which may guide the development of new treatment strategies and improve the clinical management of these diseases.

### Data selection and quality control

GWAS summary statistics for eGFR were sourced from the most extensive publicly accessible meta-analysis to date, encompassing two primary datasets (n = 1,004,040): (i) from the CKDGen consortium and (ii) from the UK Biobank.^15^ Similarly, we selected six major CVDs from large meta-analyses for greater statistical power and clinical relevance. We extracted GWAS summary statistics for AF from a genome-wide meta-analysis that included 60,620 cases and 970,216 controls of European ancestry across six studies: The Nord-Trøndelag Health Study (HUNT), deCODE, the Michigan Genomics Initiative (MGI), DiscovEHR, UK Biobank, and the AFGen Consortium.^16^ GWAS summary statistics for CAD were from a meta-analysis encompassed 181,522 cases and 984,168 controls of European ancestry, amalgamating data from nine studies not previously included along with data from the UK Biobank and CARDIoGRAMplusC4D consortium.^17^ GWAS summary statistics for VTE were obtained from a meta-analysis of seven cohorts, including the Copenhagen Hospital Biobank Cardiovascular Disease Cohort (CHB-CVDC), Danish Blood Donor Study (DBDS), Intermountain Healthcare, deCODE, UK Biobank, Million Veteran Program (MVP), and FinnGen, totaling 81,190 cases and 1,419,671 controls.^18^ GWAS summary statistics for HF included data from 47,309 cases and 930,014 controls across 26 studies from the Heart Failure Molecular Epidemiology for Therapeutic Targets (HERMES) Consortium.^19^ GWAS summary statistics for PAD involved 12,086 cases and 499,548 controls from a genome-wide meta-analysis comprising 11 independent studies.^20^ GWAS summary statistics for Stroke were sourced from a meta-analysis by the GIGASTROKE consortium, which included 73,652 patients and 1,234,808 control individuals.^21^ All summary statistics are based on individuals of European ancestry due to the limited availability of well-powered GWAS for other ancestries. Further details about the traits are available in eTable 1 in Supplement 1.

Before further analysis, we implemented rigorous quality control measures on these GWAS summary statistics by aligning them with the 1000 Genomes Project Phase 3 European population based on the hg19 genome build, removing SNPs lacking rsIDs or with duplicate rsIDs, restricting the analysis to autosomal chromosomes, and retaining only SNPs with a minor allele frequency (MAF) greater than 0.01. A total of 6,907,393 SNPs were included in the final analysis.

### Genetic correlation between eGFR and CVDs

In order to investigate the possible shared genetic basis of eGFR and CVDs, we estimated SNP-based heritability (*h^2^_SNP_*) and evaluated genome-wide genetic correlation (*r_g_*) by means of cross-trait linkage disequilibrium (LD) score regression (LDSC) method.^22^ LDSC estimates the genetic contribution to complex diseases and traits by quantifying the LD between each SNP and its neighbors. Initially, we performed univariate LDSC to estimate *h^2^_SNP_*, which reflects the proportion of phenotypic variation in a trait explained by shared genetic variants. LD scores were calculated for each SNP using genotypes of common SNPs within a 10 Mb window sourced from the 1000 Genomes Project Phase 3 European population. SNPs in the major histocompatibility complex (MHC) region (chromosome 6: 25-35 Mb) were excluded due to their intricate LD structure. We then conducted bivariate LDSC to assess the *r_g_* between eGFR and CVDs, using regression of LD scores against the product of z statistics from the respective diseases and traits. The *r_g_* ranges from -1, representing a complete negative correlation, to +1, representing a complete positive correlation, with values closer to these extremes indicating stronger correlations. The intercept from LDSC may indicate a possible sample overlap among the GWAS data sets; however, it is possible to estimate the *r_g_* in an unbiased manner, even with overlapping samples. After evaluating all pairwise *r_g_*, we applied a Bonferroni correction for multiple comparisons, setting the significance threshold at *P* < 8.33×10^-3^ (0.05 / 6).

In order to evaluate the biological basis of the shared genetic predisposition to eGFR and CVDs, we performed stratified LDSC applied to specifically expressed genes (LDSC-SEG) to identify possible enrichment of related tissue and cell types. This method integrates multi-tissue gene expression data from sources like the Genotype-Tissue Expression (GTEx) and Franke Lab, as well as multi-tissue chromatin information from the Roadmap Epigenomics and ENCODE datasets. The GTEx project provided statistics representing key tissue types and their specific expression across 49 tissues using baseline models and complete genomes. Additionally, we incorporated tissue-specific histone marker annotations from the Roadmap Epigenomics project. This included narrow peaks for DNase Hypersensitivity, H3K27ac, H3K4me1, H3K4me3, H3K9ac, and H3K36me3 chromatid. For identified relevant tissues or cell types, we used the false discovery rate (FDR) method to correct for each dataset, with a significance threshold of FDR < 0.05.

### Genetic overlap between eGFR and CVDs

LDSC primarily measures the average genetic correlation of effect sizes for all SNPs across the genome. This approach can potentially obscure the shared genetic architectures involving a mixture of concordant and discordant effect directions and may fail to capture specific overlapping loci and relevant genes. To address this limitation and quantify the polygenic overlap between eGFR and CVDs, we employed the causal mixture modeling approach (MiXeR).^23^ Firstly, univariate MiXeR analyses were performed to estimate polygenicity (i.e., the count of variants explaining 90% of *h^2^_SNP_*) and discoverability (the proportion of phenotypic variance attributable to causal variant effect sizes).^23^ We used data from the 1000 Genomes Project Phase 3 European population for our analyses and excluded SNPs within the structurally complex MHC region (CHR 6: 26-35 Mb). Subsequently, bivariate MiXeR analyses were conducted to ascertain the total count of trait-specific and shared causal SNPs visually represented in Venn diagrams. This delineation covers four components: unique causal variants for trait 1, unique for trait 2, shared causal variants, and shared non-causal variants. MiXeR calculates Dice coefficient scores (ranging from 0 to 1) to gauge polygenic overlap and estimates the proportion of SNPs with concordant effects within the shared genetic component. Additionally, MiXeR computes the *r_g_* and the correlation of effect sizes within the shared genetic component (*r_g_s*). To evaluate model fit, indicative of MiXeR’s predictive accuracy against actual GWAS data, we constructed conditional quantile-quantile (Q-Q) plots, used the Akaike Information Criterion (AIC), and generated log-likelihood plots. A model-based Q-Q plot that closely aligns with the actual Q-Q plot indicates strong predictive capability. A positive AIC value suggests the model adequately distinguishes itself from comparative models, indicating a reliable fit. Conversely, a negative AIC implies that the MiXeR model does not significantly differentiate from scenarios of maximum or minimum genetic overlap, rendering the genetic overlap estimate potentially unreliable.

### Local genetic correlation between eGFR and CVDs

To estimate local genetic correlations (local-*r_g_s*) between eGFR and CVDs within specific genomic regions and identify loci with mixed effect directions, we utilized the Local Analysis of [co]Variant Annotation (LAVA).^24^ LAVA captures detailed patterns of shared genetic loci across individuals, revealing mixed effect directions obscured by genome-wide *r_g_* estimates due to opposing effects across genomic regions. Unlike the genome-wide correlation analysis using LDSC, which considers the entire genome, LAVA divides the genome into 2,495 local LD blocks and estimates *r_g_* within each block. Differing from MiXeR’s method of estimating the proportion of shared ‘causal’ variants with concordant effects, LAVA identifies mixed effect directions by capturing significantly correlated genetic loci. Our study began with a univariate LAVA analysis to estimate local heritability for each phenotype, considering loci with *p*-values < 1×10^-4^ as significant genetic signals. This approach identified 425 loci for subsequent bivariate tests to estimate pairwise bivariate local-*r_g_s* across the genome. We adjusted the p-values for local-*r_g_s* based on the number of bivariate tests, setting a Bonferroni-corrected significance threshold at *P* < 1.18×10^-4^ (0.05/425).

Additionally, we employed Hypothesis Prioritisation for multi-trait Colocalization (HyPrColoc) on genomic regions with shared risk loci across multiple phenotypes to elucidate potential biological mechanisms linking traits. HyPrColoc, a Bayesian approach, identifies clusters of co-localization and candidate cause variants in the same genomic locus. It estimates the posterior probability (PP) of colocalization of multiple traits within a single causal variant, considering PP > 0.7 as significant evidence of colocalization. This method enhances our understanding of how genetic variants contribute to multiple traits, offering insights into complex trait interactions.

### Mendelian randomization analysis between eGFR and CVDs

The genetic correlation and overlap analysis explored the shared genetic foundation between eGFR and CVDs; however, whether their relationship is mediated by vertical or horizontal pleiotropy remains unclear. Latent Heritable Confounder Mendelian Randomization (LHC-MR)^25^ provides valuable insights into the causal relationship between eGFR and CVDs by utilizing vertical pleiotropy. This approach makes full use of all genome-wide variations, improves statistical capacity, and corrects for sample overlap, rather than just significant loci throughout the genome. Critically, LHC-MR is able to distinguish between SNPs according to their co-association with a set of traits, and to differentiate heritable confounding that results in *r_g_*. This capability allows LHC-MR to provide concurrent unbiased estimates of bidirectional causal and confounder effects. The LHC-MR framework accommodates multiple pathways through which SNPs can affect the traits, including allowing for null effects, thereby enabling precise causal effect estimations. Causal estimates from LHC-MR are presented as odds ratios (ORs) with corresponding 95% confidence intervals (CIs). Causality is considered unidirectional if *P* is lower than the Bonferroni-corrected threshold (0.05 / 6 / 2 = 4.17×10^-3^) and the *P*-value of the effect in the opposite direction is greater than 0.05. Bidirectional causality is considered if *P* < 4.17×10^-3^ in both directions. Additionally, we employed several methods as sensitivity analyses to validate the results, including the inverse variance weighted (IVW) method, the weighted median, MR-Egger, simple mode, and weighted mode. These analyses help confirm the robustness of the causal inferences drawn from our study.

## SNP-level analysis

### Identification of pleiotropic loci between eGFR and CVDs

In order to explore the effect of horizontal pleiotropy between eGFR and CVDs, we used Pleiotropic Analysis under the Composite Null Hypothesis (PLACO) to clarify the shared genetic mechanisms underlying these conditions.^26^ PLACO identifies pleiotropic loci between two traits by testing the compound null hypothesis that a locus is related to either none or only one of them. This hypothesis is further subdivided into specific sub-scenarios: (1) M00, indicating no association with any trait; (2) M10 and M01, each indicating an association with only one of the traits; and (3) M11, representing a pleiotropic association with both traits. This method calculates the PLACO statistics by multiplying the Z-scores of the two traits (Z_trait1_ × Z_trait2_) while excluding SNPs with squared Z-values above 100 to mitigate spurious signals of pleiotropy. SNPs are considered to exhibit significant genome-wide pleiotropy if their *P_PLACO_*is less than 5×10^-8^.

### Genomic loci definition and functional analysis

To identify pleiotropic SNPs associated with eGFR and CVDs, we utilized the Functional Mapping and Annotation of Genome-Wide Association Studies (FUMA) platform. FUMA integrates data from a variety of biological databases to improve functional annotation, gene prioritization, and interactive visualization of GWAS outcomes.^27^ We calculated LD scores using the 1000 Genomes Phase III European population as a reference. Firstly, independently significant SNPs achieving genome-wide significance (*P* < 5.0 × 10^−8^ and r^2^ < 0.6) were identified. Lead SNPs were then determined based on their independence (r^2^ < 0.1). Genomic risk sites were delineated by merging lead SNPs within 500 kb of each other, allowing these sites to contain multiple lead SNPs. The top lead SNP at each site, defined by the lowest P value, was selected. Additionally, the directional impact of these sites was assessed by comparing Z-scores between eGFR and CVD, providing insights into their potential roles in these conditions. A GWAS site was considered novel if none of its lead, independent, or candidate SNPs overlapped with SNPs previously reported in GWAS meta-analyses. Using LocusZoom, we created a regional association plot with gene tracks, allowing us to examine the details of the relationship between each locus. To predict the functional outcomes of Top SNPs, we matched SNPs to databases containing established functional annotations, including the Annotate Variation (ANNOVAR) category, Combined Annotation-Dependent Depletion (CADD) score, RegulomeDB (RDB) score, and chromatin state.^28–30^ The CADD score is used to predict the deleteriousness of SNP effects, incorporating 63 functional annotations. A threshold of 12.37 is generally recognized as indicative of deleterious variants, and thus, we filtered SNPs with a CADD score greater than 12.37 for location mapping. In order to prioritize eQTL genes and explore their potential regulatory role, we used RegulomeDB, which provides functional interpretation of SNPs based on curated references. The RegulomeDB scoring system, ranging from 1a (most regulatory potential) to 7 (least), helps evaluate the regulatory likelihood of SNPs. Additionally, chromatin states were used to delineate the regulatory landscape of genomic regions, with 15 classes of states predicted using 5 chromatin markers across 127 epigenomes via the ChromHMM tool.^31^ Two methods were employed to map SNPs to genes: (i) positional mapping, which assigns SNPs to genes based on their physical proximity (within a 10kb window) to known protein-coding genes in the human reference assembly, and (ii) eQTL mapping, which links SNPs to genes based on significant eQTL associations, where allelic variations in an SNP correlate with variations in gene expression levels, as identified using the GTEx database. This approach improves our understanding of the genetic structure by linking SNPs to potential functional outcomes.

### Colocalization analysis

For FUMA-annotated pleiotropic loci, we performed a Bayesian colocalization analysis to identify potential common causal variations between trait pairs. COLOC employs a Bayesian framework to calculate the PP of five distinct hypotheses concerning shared causal variation within a genomic region.^32^ These hypotheses assess whether one or both traits share causal variations at a specific locus. Specifically, the hypotheses are as follows: i) PPH0: Neither trait has causal variation at the locus; ii) PPH1: Only the first trait has causal variation at the locus; iii) PPH2: Only the second trait has causal variation at the locus; iv) PPH3: Each trait has a different causal variation within the locus; v) PPH4: Both traits share a causal variation within the locus.^33^ The SNP exhibiting the highest PPH4 within the locus is identified as the candidate causal variant. Loci are considered colocalized if the PPH4 exceeds 0.7, indicating a strong likelihood that the locus harbors shared causal variations for the traits under study.

### Gene-level analysis using MAGMA, eMAGMA and TWAS

To identify candidate pleiotropic genes further, we conducted Multi-marker Analysis of GenoMic Annotation (MAGMA) based on results from PLACO and individual GWAS. MAGMA uses the SNP average model within the multi-regression framework to derive *P* values, and to estimate the association of the gene, and to adjust for factors such as the size of the gene, the number of SNP for each gene, and the LD between markers.^34^

Our analysis referenced the 1000 Genomes Project Phase 3 European population and used the Genome Reference Consortium Human Build 37 (hg19) for SNP locations and gene annotations. We analyzed 17,636 protein-coding genes on the MAGMA software website, focusing on genes containing at least ten SNPs to ensure computational stability. Gene based testing involves extending 10kb upstream and downstream of the gene transcription start and end sites. Because of their complicated LD patterns, we ruled out MHC regions (chr6: 25 - 35 Mb). Strict Bonferroni correction was used for adjustment for multiple trials, and in the MAGMA analysis based on the PLACO results, a significant threshold of *P* < 4.23×10^-7^ (0.05 / 17636 / 6) was established.

The inherent limitations of MAGMA, which assigns SNPs based solely on proximity to genes without accounting for functional associations such as gene regulation, can hinder its effectiveness in elucidating the underlying mechanisms of genetic variants. To overcome this and delve deeper into the functional implications of genetic variants associated with eGFR and CVDs, we employed the EQTL-informed MAGMA (E-MAGMA).^35^ Operating within the same statistical framework as MAGMA, E-MAGMA uses a multi-principal component linear regression model to leverage tissue-specific eQTL data from multiple sources, aiming to identify potential causal genes for phenotypic traits. Our study utilized eQTL data from 47 tissues, enriched for differentially expressed genes in tissues as provided by the Genotype-Tissue Expression Project version 8 (GTEx v8), and referenced the 1000 Genomes Project Phase 3 European population. To mitigate potential confounding effects from the broad range of tissues, we focused on 11 specific tissues that were identified by LDSC-SEG analysis, including arterial, adipose, cardiac, whole blood, liver, EBV-transformed lymphocytes, and kidney tissues. We excluded kidney tissue from our analysis due to the absence of reference in E-MAGMA. We applied Bonferroni-adjusted *p*-value thresholds to accommodate multiple testing, calculated based on the number of genes analyzed in E-MAGMA and the number of statistically significant trait pairs tested. For example, the threshold for subcutaneous fat was defined as *P* < 8.67 × 10^-7^ (0.05 / 9,603/ 6).

We conducted the Transcriptome-wide Association Study (TWAS) using single-trait GWAS results to explore the tissue specific gene expression correlation between eGFR and CVDs. TWAS combines eQTL and summary association statistics from large scale GWAS to identify genes that are associated with complex traits.^36^ For gene expression prediction, we utilized the FUSION tool with default settings, employing various methods, including best linear unbiased prediction (BLUP), Bayesian sparse linear mixed model (BSLMM), least absolute shrinkage and selection operator (LASSO), elastic net (ENET), and Top SNP, which were used to estimate the cis-genetic component of tissue-specific gene expression. We selected the most effective predictive model to determine gene expression weights and used GWAS summary statistics to conduct the TWAS. In addition, we used the Bonferroni correction to adjust for multiple comparisons between the different tissue types analyzed.

### Pathway-level analysis using MAGMA and Metascape

We conducted a MAGMA gene set analysis to elucidate the biological functions of genes exhibiting pleiotropic effects on eGFR and CVDs. This analysis employs a competitive gene set framework to assess whether specific gene sets are more strongly associated with a particular phenotype than the broader genomic background. Our approach integrates gene definitions and their respective signals using MAGMA’s gene-based multi-marker method. The analyzed gene set is derived from Gene Ontology (GO) and Reactome pathways, as listed in the Molecular Feature Database (MSigDB v7.5). The significance threshold was set to *P* < 0.05 / (7744 + 1654) / 6, adjusted for the number of GO and Reactome pathways, as well as the number of trait pairs analysed. To elucidate the biological processes and signaling pathways associated with eGFR and CVDs, we conducted Metascape analysis on the overlapping genes identified by MAGMA and EMAGMA. Metascape provides comprehensive annotation of gene and protein, enrichment analysis, and protein-protein interaction networks, which can help to better understand the functions of genes.^37^ We used Metascape to perform GO annotation and Reactome enrichment analysis, which utilizes the hypergeometric test to identify significant ontology terms. The GO resource offers a framework and concepts to describe gene product functions across all organisms, while the Reactome knowledgebase details cellular processes, such as signaling, transport, DNA replication, and metabolism, as ordered networks of molecular transformations. Pathways with a *p*-value < 0.01 were considered significant.

### Proteome-wide Mendelian Randomization analysis using SMR

To investigate potential associations between plasma protein levels and disease susceptibility, we employed Summary data-based Mendelian Randomization (SMR). This analysis integrated plasma protein quantitative trait locus (pQTL) summary statistics from the UK Biobank Pharmaceutical Proteomics Project (UKB-PPP) with GWAS summary statistics for various diseases. The UKB-PPP data, derived from the Olink proteomics platform, includes genetic associations for 2,940 plasma proteins across a cohort of 34,557 Europeans. In this context, cis-pQTLs are defined as SNPs located in the 1Mb window surrounding the transcription start site (TSS) of each protein. For SMR analysis, we only took into account the cis-pQTLs related to plasma protein levels at the genome-wide significance threshold (*P* < 5 × 10^-8^). SMR, a method that utilizes summary-level data, assesses potential causal relationships between exposures (e.g., plasma protein levels) and outcomes (e.g., traits or diseases). In order to distinguish between pleiotropy and linkage in the cis-pQTL region, we tested multiple SNPs using the heterogeneity of instrument-dependent (HEIDI) method. Results indicating pleiotropy (HEIDI test *P* < 0.01) were excluded from further analysis. In recognition of the limitations of single-SNP analyses, we also performed a sensitivity analysis using a multi-SNP-SMR test, with a P value of < 0.05 being significant. A Bonferroni correction was applied to the number of unique proteins to be analyzed, and a significance threshold was set at *P* < 3.65×10^-6^ (0.05 / 1958 / 7). Additionally, we performed COLOC analysis to determine if the same causal variation is responsible for the association of protein levels with disease phenotype. The PP.H4 > 0.7 for shared causal variants indicates significant colocalization of GWAS and pQTL.

## Result

### Genome-wide genetic correlation between eGFR and CVDs

After implementing stringent quality control, we utilized LDSC to assess *h^2^_SNP_* and genome-wide *r_g_* between eGFR and CVDs. We used univariate LDSC to estimate *h*^2^ for eGFR and each CVD. The SNP heritability estimate was highest for eGFR (*h^2^_SNP_* = 0.0708, SE = 0.0036). Among these CVDs, heritability estimates ranged from 0.0060 for Stroke, the lowest (SE = 0.0005), to 0.0324 for CAD, the highest (SE = 0.0019) (Supplementary Fig. 1a and Supplementary Table 2a).We then conducted bivariate LDSC analysis to explore the *r_g_* between eGFR and CVDs. This revealed significant negative *r_g_* for two trait pairs: eGFR and Stroke (*r_g_* = -0.0985, SE = 0.0309) and eGFR and VTE (*r_g_* = -0.0783, SE = 0.0238), both surpassing the Bonferroni-corrected significance threshold (*P* < 8.33×10^-3^) (Supplementary Fig. 1b and Supplementary Table 2b). However, no significant *r_g_* was found between eGFR and AF, CAD, HF, or PAD.

### Polygenic overlap and local genetic correlation between eGFR and CVDs

Nevertheless, previously described *r_g_* may underestimate the genetic overlap between eGFR and CVDs. The fact that there is no significant *r_g_* does not necessarily mean that there is no common genetic component among these traits. Indeed, the *r_g_* measure does not distinguish between mixtures of concordant and discordant genetic effects and a true absence of genetic overlap. To address this limitation and more comprehensively elucidate the shared genetic underpinnings of complex polygenic features, we employed advanced analytical tools, MiXeR and LAVA, specifically designed to provide more detailed insights into genetic architectures that traditional analyses might obscure.

We conducted MiXeR analyses to quantify the polygenic genetic overlap between eGFR and CVDs, accounting for mixed effect directions. Initially, univariate MiXeR analyses estimated that approximately 2,044 variants (SD = 111) influence eGFR. Among the CVDs, HF had the highest polygenicity, with 2,286 ’causal’ variants accounting for 90% of HF *h*^2^ (SD = 215), followed by CAD with 1,431 variants (SD = 311), and stroke with 1,039 variants (SD = 120) (Supplementary Table 3a). Subsequently, bivariate analyses revealed significant polygenic overlap between eGFR and these CVDs, with Dice coefficients ranging from 0.024 to 0.457 (Fig. 2a, Supplementary Fig. 2, Supplementary Table 3b). Notably, eGFR displayed a moderate genetic overlap with diseases like CAD (Dice = 0.457), HF (Dice = 0.39), and stroke (Dice = 0.35). Among these, the eGFR-HF pair possessed the highest number of shared ’causal’ variants (841, SD=151), accounting for substantial proportions of variants affecting each condition (41.1% for eGFR and 36.8% for HF), suggesting highly similar genetic architectures between these two traits. However, the genetic correlation (4.78×10^-4^, SD=9.83E-3) and the correlation coefficients of genetic risk (9.62×10^-4^, SD=0.024) between eGFR and HF were notably weak. This extensive genetic overlap, coupled with weak *r_g_*, highlights the existence of mixed effects, as demonstrated by the proportion of shared ’causal’ variants with consistent effects (0.50, SD = 0.007). A similar relationship was observed between eGFR-CAD and eGFR-Stroke, with the *r_g_* and *r_g_s* (*r_g_* = -0.098; *r_g_s* = -0.269) of eGFR-Stroke consistent with their LDSC-estimated significant negative genetic correlations. Compared to diseases with lower polygenicity, such as AF, VTE, and PAD, eGFR is characterized as a highly polygenic trait, leading to significant disparities in the number of shared and unique ’causal’ variants. For instance, the eGFR-VTE pair demonstrated mild genetic overlap (Dice = 0.177) with 211 shared variants (SD = 83), accounting for 10.3% of eGFR’s and 66.2% of VTE’s heritability, and exhibited the highest genetic risk correlation (*r_g_*= -0.115, SD = 0.007). Conversely, the eGFR-PAD pair had the fewest shared variants (28, SD = 15), but the shared genetic components showed the strongest correlation (*r_g_s* = 0.567, SD =0.351), indicating that while many eGFR-associated variants do not influence PAD, those that do have highly similar effects (Supplementary Fig. 2).

**Figure 2:**
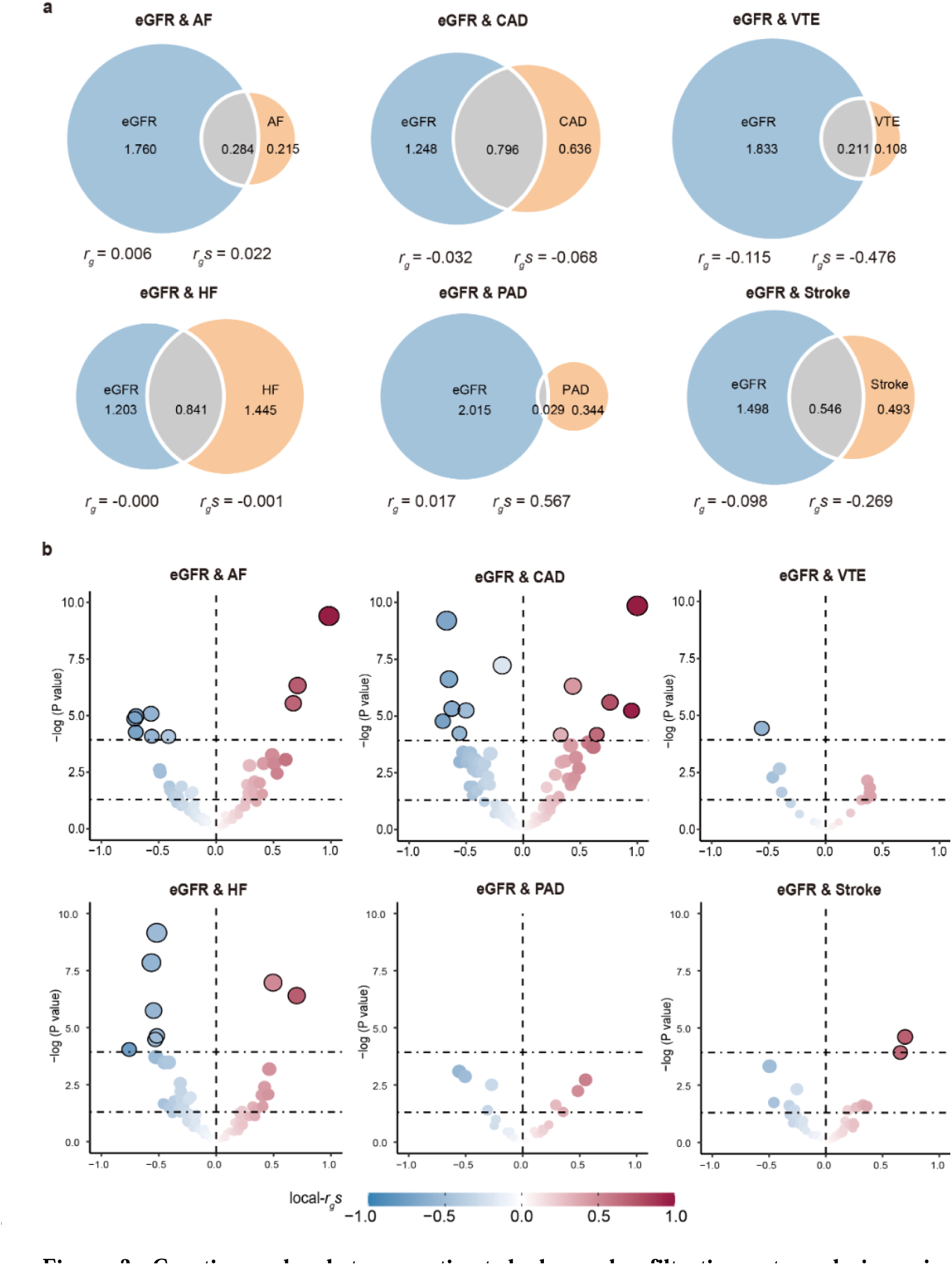
Genetic overlap between estimated glomerular filtration rate and six major cardiovascular diseases exceeds genome-wide genetic correlation. Genetic overlap and local genetic correlation between eGFR and six major CVDs investigated by MiXeR and LAVA. a: MiXeR Venn diagrams showing common and unique effect trait variants showing polygenic overlap (grey) between eGFR (blue) and CVDs (orange). Numbers in the Venn diagrams indicate the estimated number of common and unique effect trait variants (in thousands) that explain 90% of the SNP heritability and standard error in each phenotype. Circle size indicates the degree of polygenicity for each trait, with larger circles indicating stronger polygenicity. We also provide genome-wide genetic correlation (*r_g_*) and genetic correlation of shared variants (*r_g_s*). b: LAVA volcano plots showing local genetic correlation coefficients (local-*r_g_s*, y-axis) for eGFR and CVDs with -log10 *p*-values for each trait pair of analyses for each locus. Loci above the horizontal line are significant at *P* < 0.05 (negative correlation for blue dots, positive correlation for red dots). Larger points with black circles indicate loci significantly associated after Bonferroni correction (*P* < 0.05 / 425). LAVA-estimated local-*r_g_s* is shown on the blue-red scale. eGFR, estimated glomerular filtration rate; AF, atrial fibrillation; CAD, coronary artery disease; VTE, venous thromboembolism; HF, heart failure; PAD, peripheral arterial disease.

LAVA, which calculates local-*r_g_s*, was employed to explore further the relationships between eGFR and CVDs, revealing mixed effect directions. First, univariate analyses were conducted to identify heritable regions, filtering out loci devoid of univariate heritability (Supplementary Table 4), leading to 425 bivariate tests. While no significant genome-wide *r_g_*emerged between eGFR and diseases such as AF, CAD, HF, or PAD, LAVA identified specific regions with significant local-*r_g_s* (*P* < 0.05, Fig.2b and Supplementary Table 5). This was particularly notable between eGFR and CAD, where 27 positively and 28 negatively correlated regions were found, suggesting that the overall non-significant genome-wide correlation could result from counterbalancing local effects. Similarly, mixed effects were observed with other diseases: eGFR and AF exhibited 17 positive and 18 negative regions, HF showed 5 positive and 4 negative regions, and PAD had an equal number of positive and negative regions (4 each). Contrary to these, eGFR and VTE displayed more negatively correlated regions (12 positive / 23 negative), aligning with the negative genome-wide correlations estimated by LDSC. However, the findings for eGFR and Stroke, which presented more positively than negatively correlated regions, did not consistently align with the negative *r_g_* estimated by LDSC, though 5 negatively correlated regions were still identified. After applying the Bonferroni correction, 36 genetic regions showed significant local-*r_g_s*. Notably, three genetic regions were linked to multiple trait pairs: LD block 96 on chromosome 1 was associated with both eGFR-CAD and eGFR-VTE; LD blocks 673 on chromosome 4 and 1057 on chromosome 6 exhibited correlations between eGFR-AF and eGFR-CAD. Further analysis using HyPrColoc identified shared causal variants, with SNP rs12740374 in LD block 96 showing strong co-localization evidence of influencing eGFR-VTE and CAD with a PP greater than 0.7.

In summary, we systematically quantified the genetic overlap between eGFR and six major CVDs using MiexR and LAVA, going beyond simple genome-wide *r_g_*. These analyses highlighted significant shared genetic bases between eGFR and the CVDs, which were not apparent from traditional genome-wide studies alone. Building on these findings, we performed further pleiotropy analysis to delve deeper into the molecular mechanisms underlying these trait pairs.

### Causal effects identified between eGFR and CVDs

Based on the above-mentioned common genetic basis between eGFR and CVDs, we implemented multiple pleiotropy methods to reveal the potential mechanisms of vertical or horizontal pleiotropy. Among them, in terms of vertical pleiotropy, we used LHC-MR technology to study causal relationships while controlling bias and confounding factors. Our analysis found that AF had a significant reverse causal effect on eGFR (OR: 0.806; 95% CI = 0.720 to 0.903), which suggests that the presence of AF may reduce eGFR. In addition, we observed another pair of significant negative causal relationships, indicating that an increase in eGFR would reduce the risk of VTE (OR = 0.931; 95% CI = 0.899 to 0.964) (Fig. 3, Supplementary Table 6a). These findings were confirmed by the results of the IVW method. In summary, our MR study provides strong evidence for the causal relationship between AF and eGFR and between eGFR and VTE (Supplementary Fig. 5, Supplementary Table 6b). However, it must be recognized that vertical pleiotropy alone cannot fully explain the common genetic basis between eGFR and CVDs.

**Figure 3:**
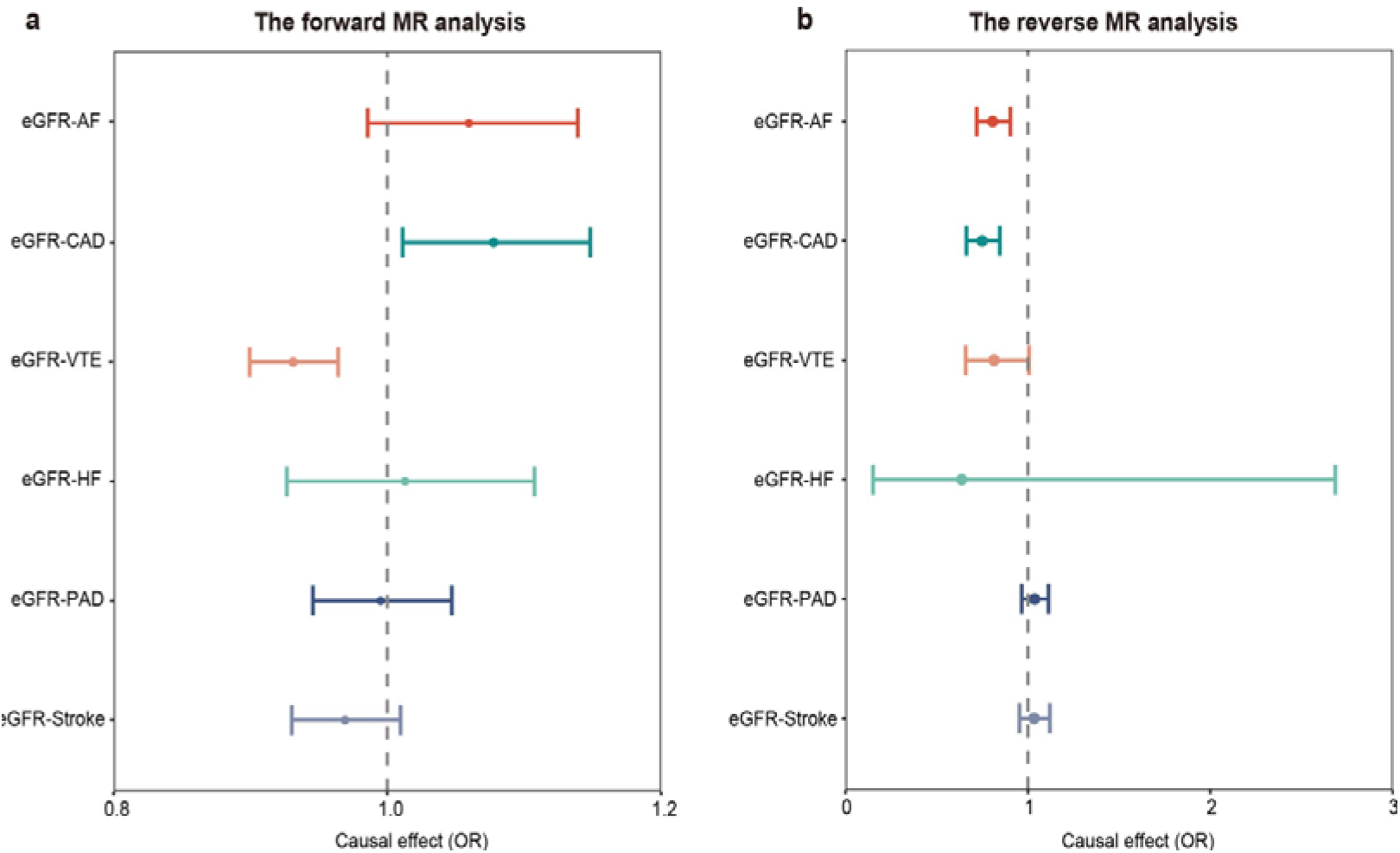
Inference of causal relationship between estimated glomerular filtration rate and six major cardiovascular diseases. Forest plots of the causal relationship between eGFR and six major CVDs using the LHC-MR method. a: indicates the estimated causal relationship of eGFR on CVDs. b: indicates the estimated causal relationship of CVDs on eGFR. Circles indicate odds ratio (OR) estimates, and error bars indicate 95% confidence intervals. OR > 1 indicates a positive association, and OR < 1 indicates a negative association. eGFR, estimated glomerular filtration rate; AF, atrial fibrillation; CAD, coronary artery disease; VTE, venous thromboembolism; HF, heart failure; PAD, peripheral arterial disease.

### Pleiotropic genomic loci identified between eGFR and CVDs

Given the extensive genome-wide genetic overlap, we further tested pleiotropic SNPs between eGFR and CVDs by PLACO, and a total of 20,073 SNPs (*P* < 5×10^-8^) were identified as significant pleiotropic variants. FUMA aggregated these SNPs into 508 loci, involving 226 unique chromosomal regions, of which 32 loci appeared in over 50% of trait pairs (Fig. 4, Supplementary Fig 6, Supplementary Table 7). Notably, loci such as 6q25.3 (SLC22A1), 8p23.1 (BLK), 9q34.2 (ABO), and 6p21.1 (*CRIP3*) showed overlap across all six trait pairs, while 12q24.12 (*ALDH2*) appeared in five. These broad pleiotropic effects are exemplified by the common variants at 6p21.1, which are associated with both polycystic kidney disease and stroke, and by 12q24.12, a locus significant for CAD and stroke. Interestingly, 24 pleiotropic loci had not been reported in previous eGFR and CVD studies, which reported 81 and 364 loci, respectively.

**Figure 4:**
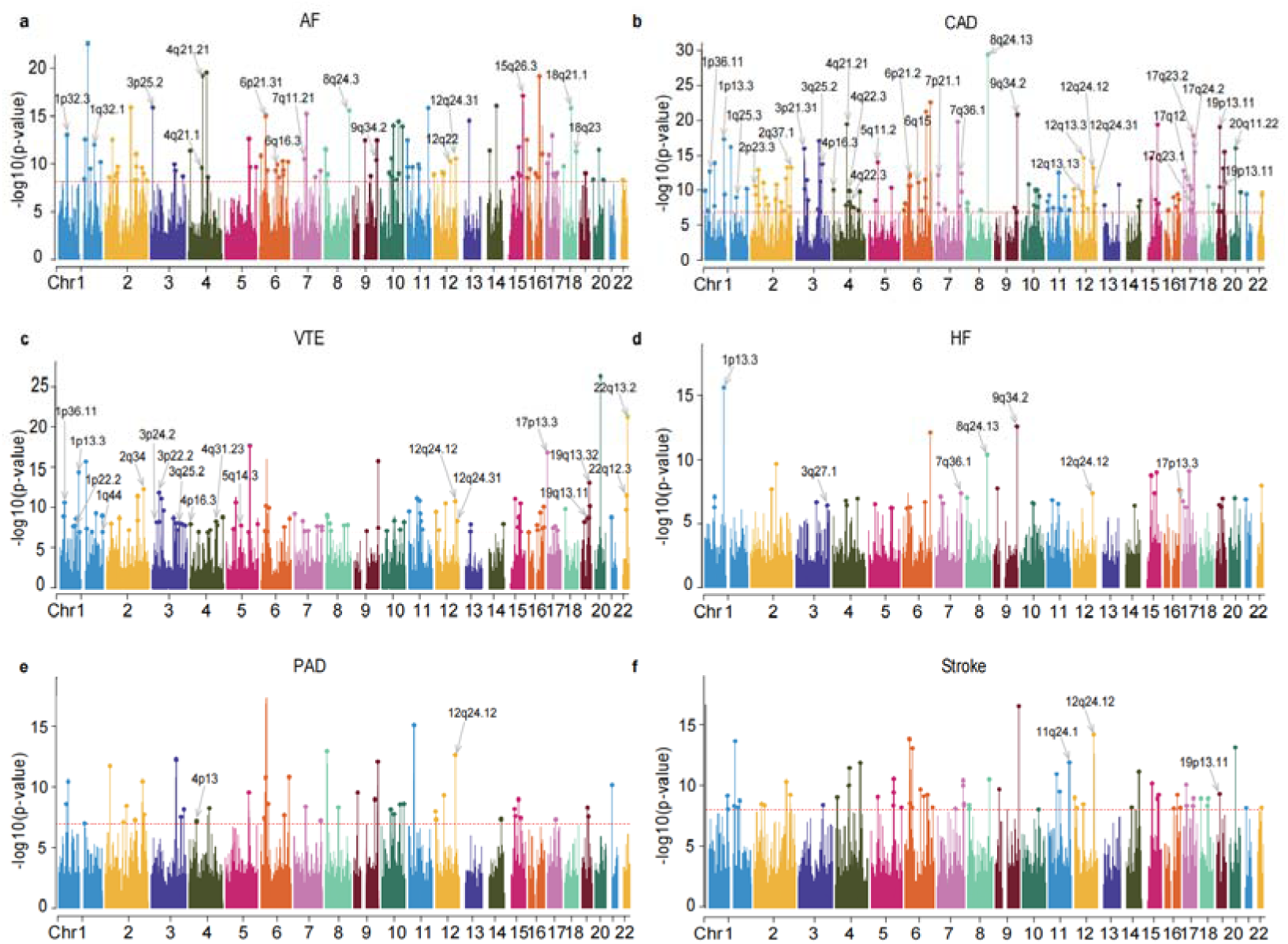
Manhattan plot of PLACO results for estimated glomerular filtration rate and six major cardiovascular diseases. Manhattan plots reflect chromosomal position (x-axis) and negative log10-transformed *P*-values (y-axis) for each SNP. Horizontal lines indicate genome-wide significant P-values -log10 (5×10^-8^). The r^2^ threshold for defining independent significant SNPs was set to 0.2, and the maximum distance between LD blocks merged into one locus was set to 500 kb. *P*-values were derived using GWAS multi-trait analysis in discovery studies, and independent genome-wide significant associations with the smallest *P*-values (top SNPs) are circled in colored circles. Only SNPs that were common in all summary statistics were included. Labels are chromosomal regions where genomic risk loci with strong colocalization evidence (PP.H4 > 0.7) are located. eGFR, estimated glomerular filtration rate; AF, atrial fibrillation; CAD, coronary artery disease; VTE, venous thromboembolism; HF, heart failure; PAD, peripheral arterial disease.

Targeted analysis within the 508 pleiotropic loci revealed that 139 top SNPs (27.4%) indicated an increased risk for these traits, while 118 (23.2%) indicated a reduced risk. The remaining 251 top SNPs (49.4%) showed discordant associations, highlighting potential contrasting biological mechanisms. Functional annotation using FUMA identified that 330 SNPs (65.0%) were intronic variants, 127 (25.0%) were intergenic, and 21 (4.1%) were exonic, including 14 mRNA and 7 non-coding RNA exon variants. Moreover, 40 SNPs in these risk loci, including rs34312154 with the highest CADD score of 31, may be potentially harmful, affecting genes like *HOXB1* to *HOXB8*. Additionally, 58 SNPs identified by RegulomeDB as potentially affecting transcription factor binding underscore the regulatory significance of these findings, with rs4918003 showing the most potent evidence of regulatory potential (RDB: 1b).

Further analysis of co-localization at 508 identified loci revealed that 74 loci (14.6%) exhibit high PP (PPH4 > 0.7), indicating strong evidence of shared genetic signals (Fig. 4, Supplementary Fig. 7, Supplementary Table 7). Of these, 71 top SNPs were identified as candidate common pathogenic variants. Notably, the 1p13.3 locus showed significant pleiotropy, influencing the trait pairs of eGFR-CAD, eGFR-VTE, and eGFR-HF, with PP.H4 values ranging from 0.956 to 0.998. Subsequent HyPrColoc analysis identified rs660240 in the CELSR2 gene at this locus as exhibiting strong co-localization for both eGFR and HF and eGFR and VTE, with PP exceeding 0.7.

### Identification of pleiotropic genes between eGFR and CVDs

We used MAGMA analysis to convert the SNP level signals of identified eGFR-CVDs into gene level signals, focusing on potential pleiotropy within or overlapping 508 pleiotropic loci. This analysis identified 802 significant pleiotropic genes, with 579 being unique. Notably, 379 genes were implicated in more than one trait pair, providing robust statistical evidence for shared genetic inheritance across diverse traits (Fig. 5, Supplementary Table 11). Specific genes such as *ATXN2* (located at 12q24.12) significantly impacted five trait pairs. Additionally, genes including *ABO* (9q34.2), *BRAP* (12q24.12), *SH2B3* (12q24.12), *L3MBTL3* (6q23.1), *MMP24* (19q13.32), *MRPS21* (1q21.2), and *PRPF3* (1q21.2) were significant across four trait pairs, underscoring their pivotal roles in these genetic networks. Remarkably, the 12q24.12 locus, home to *ATXN2*, *BRAP*, and *SH2B3*, is critical in assessing genetic risks for hypertension^38^ and CKD, with *ATXN2* linked to myocardial infarction, renal function, and hypertension.^39^ Among the genes identified, 118 were not previously explored in eGFR studies, and 410 were new to CVD research, highlighting significant novel findings. Positional mapping in FUMA confirmed 97.88% of these risk genes identified by MAGMA, further solidifying the evidence for a shared genetic framework across these trait pairs (Supplementary Table 9).

**Figure 5:**
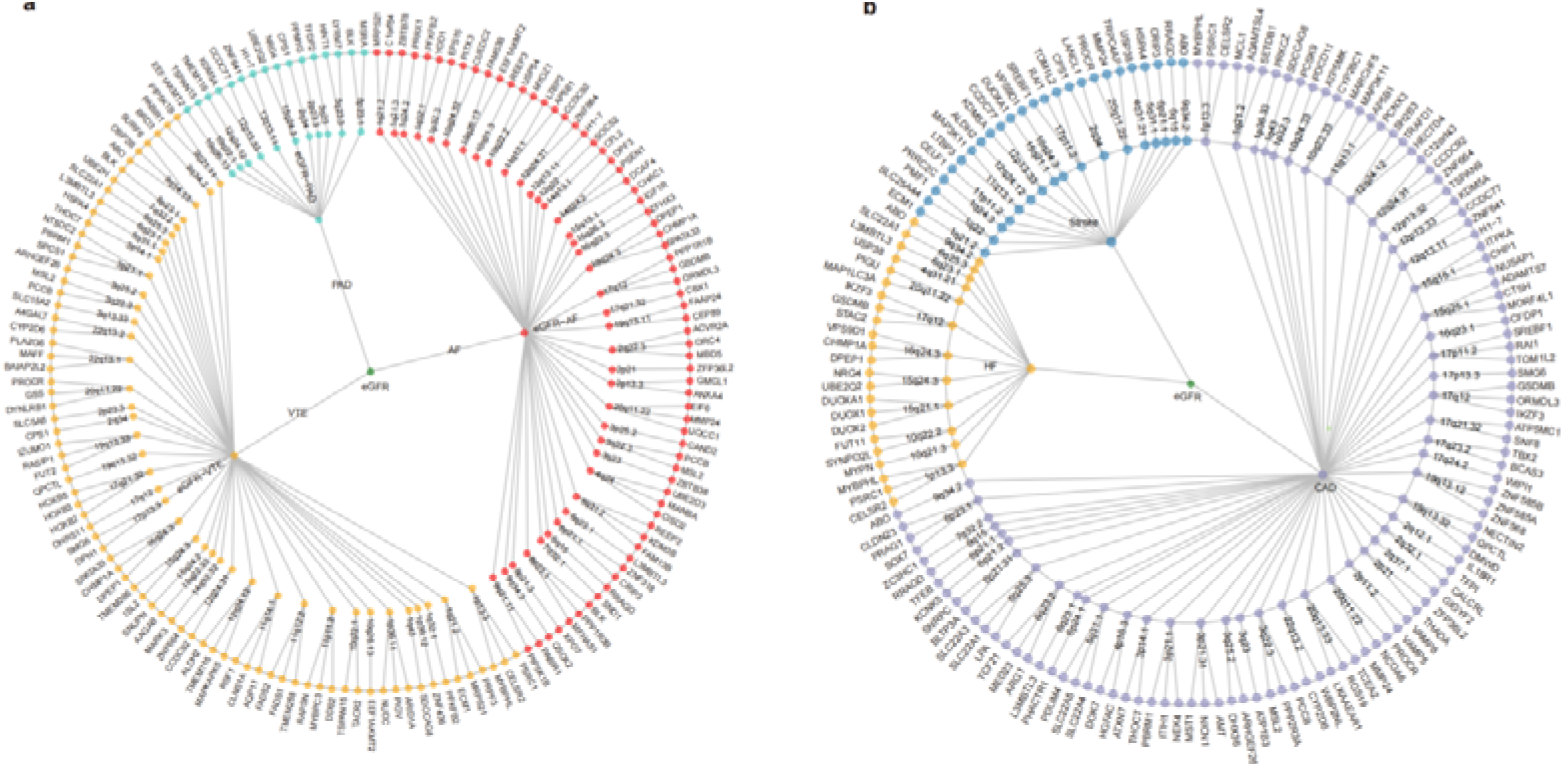
The overall situation of the pleiotropy association between estimated glomerular filtration rate and six major cardiovascular diseases. Based on the polygenic nature of the Mixer results, we separated the pleiotropic association between eGFR and CVDs into two network diagrams, showing the pleiotropic loci and genes identified in the six trait pairs. a: The circular dendrogram is centered on eGFR (inner circle) and branches out for the three CVDs (second circle, AF, VTE, and PAD). These three trait pairs show 91 pleiotropic loci (third circle) corresponding to 159 pleiotropic genes (outer circle). b: The circular dendrogram is centered on eGFR (the inner circle) and branches out for the three CVDs (second circle, CAD, HF, and stroke). Another three trait pairs show 80 pleiotropic loci (third circle) corresponding to 153 pleiotropic genes (outer circle). For trait pairs with three or more pleiotropic genes, we only show the top three pleiotropic genes according to priority, and these genes show statistical priority decay in a clockwise direction. For example, eGFR-AF-12q24.31-CCDC92 and eGFR-AF-12q24.31-ZNF664 indicate that 12q24.31 is a shared locus for this pair of specific diseases involving CCDC92 and ZNF664. eGFR, estimated glomerular filtration rate; AF, atrial fibrillation; CAD, coronary artery disease; VTE, venous thromboembolism; HF, heart failure; PAD, peripheral arterial disease.

We employed the LDSC-SEG method to perform tissue-specific genetic analysis, identifying distinct tissue associations for each trait. The analysis revealed significant gene expression enrichment for eGFR in the renal cortex and liver tissues. Similarly, AF showed considerable enrichment in the atrial appendage and cardiac left ventricular tissues, while CAD was significantly enriched in the aorta, coronary artery, and tibial artery tissues, with all exceeding an FDR threshold of 0.05. Subsequent LDSC-SEG chromatin analysis corroborated these findings, enhancing the resolution by detecting significant tissue chromatin signals associated with eGFR, AF, and CAD traits (Supplementary Fig. 3, Supplementary Table 14).

We conducted E-MAGMA analysis on ten related tissues using tissue-specific eQTL information, which identified 1,276 pleiotropic genes that remained significant after Bonferroni correction, showing strong enrichment in at least one of the analyzed tissues (Supplementary Table 15). Particularly noteworthy, 26 genes were prevalent in over 50% of trait pairs, with *KAT5* and *PROCR* appearing in five trait pairs each and *BLK* and *SLC22A1* in four. *KAT5*, or lysine acetyltransferase 5, serves a dual role; it regulates the level of acetylation of target proteins, affecting the expression of inflammation related genes and the intensity of inflammatory responses. This regulatory function is crucial, as abnormal expression or dysfunction of *KAT5* can elevate cardiovascular disease risks by affecting the inflammatory pathway.^40^ Additionally, *KAT5* acts as a DNA repair factor, vital for preventing ischemic acute kidney injury (AKI), primarily by regulating glomerular filtration.^41^ Therefore, the normal function and activity of *KAT5* are critical in preventing or alleviating AKI and indirectly maintaining or enhancing eGFR levels. We also performed the TWAS to assess the tissue specificity of pleiotropic genes for each trait, leading to the discovery of 354 new genes associated with eGFR and 142 new genes related to various CVDs (Supplementary Table 16). Furthering our investigation, we utilized FUMA for eQTL mapping, identifing 2,049 genes, of which 71.40% were confirmed by our analysis to be associated with the traits under study (Supplementary Table 9).

MAGMA and E-MAGMA analyses have jointly identified 424 pleiotropic genes, including 314 unique ones, shedding light on the common mechanisms underlying various trait pairs (Supplementary Table 11). Notably, 76 of these genes recur across multiple trait pairs, with genes such as *MMP24*, *L3MBTL3*, and *ABO* being prominent in half or more of the analyzed traits. Specific genes like *ABO* (9q34.13), *PROCR* (20q11.22), *BLK* (8p23.1), *ALDH2* (12q21.31), and *SLC22A1* (6q25.3) each featured in three different trait pairs, highlighting their significant roles in multiple pathologies. For instance, *L3MBTL3* has been implicated in renal dysfunction and cardiovascular pathogenesis, illustrating the interconnectedness of renal and cardiovascular systems. *MMP24*’s dysregulated expression, associated with renal tubular atrophy and diabetic nephropathy (DN), may contribute to structural and functional kidney impairments that exacerbate cardiovascular risk. In addition, the *ABO* gene, which is associated with changes in von Willebrand factor (VWF) plasma levels, plays a key role in regulating platelet function and atherosclerosis, thus affecting the severity of CAD. While the precise mechanisms connecting ABO genes to eGFR remain elusive, their association with conditions like type 2 diabetes (T2D) and hypertension, which are main risk factors for CKD, likely impacts eGFR.^42^

### Shared biological mechanisms between eGFR and CVDs

We employed the MAGMA algorithm to assess pathway enrichment among the identified genes, setting a stringent Bonferroni-corrected threshold (*P* < 8.87×10^-7^). This analysis revealed 78 significantly enriched pathways, comprising 73 Gene Ontology Biological Process (GO BP) terms and 5 Reactome gene set pathways, indicative of key biological processes (Supplementary Table 17a). Notably, pathways such as the positive regulation of macromolecule biosynthetic processes, transcription by RNA polymerase II, and RNA metabolic processes were enriched in half or more of the trait pairs, highlighting their crucial roles in the regulation of core biological processes that contribute to the development of the studied traits.

Further analysis of pathway enrichment for overlapping genes, identified in multiple trait pairs from MAGMA and E-MAGMA analyses, was conducted using the Metascape website (Supplementary Table 17b). This analysis pinpointed 16 significantly enriched pathways predominantly involved in regulating biosynthetic and metabolic processes, lipid metabolism, and receptor signal transduction. The pathways related to glutathione breakdown and stem cell maintenance were critical and had profound implications for eGFR and CVDs. The antioxidant and anti-inflammatory properties of glutathione are essential for cardiovascular protection. However, its degradation may elevate the risk of CVDs through mechanisms linked to renal insufficiency. Moreover, the protective roles of endothelial progenitor cells (EPC) and stem cell factor (SCF) within the kidney are critical for maintaining eGFR and CVDs health, implying that diminished activity in these pathways could adversely affect renal and cardiovascular systems.

### Proteome-wide Mendelian Randomization analysis for eGFR and CVDs

We conducted a SMR analysis utilizing GWAS summary data for eGFR, six major CVDs, and plasma protein pQTLs. This analysis identified 78 risk proteins, 32 linked to eGFR and 46 to CVDs, validated through HEIDI evaluation (*P* > 0.01), multiple SNP-SMR sensitivity analysis (*P* < 0.05), and Bonferroni corrections (*P_SMR_*< 0.05 / (1958×7)). Subsequently, we investigated potential pleiotropic effects within specific eGFR-CVD pairs, identifying three signature pairs that contain pleiotropic proteins (Supplementary Table 18). Notably, CELSR2 was common between eGFR-CAD and eGFR-VTE, while FGF5 linked eGFR-AF and eGFR-CAD. Additionally, a colocalization analysis of these 78 proteins sought to identify common pathogenic variants associated with specific eGFR or CVDs signatures. This analysis showed strong evidence of colocalization for 49 proteins (41 unique), with 17 proteins associated with eGFR and 32 with CVDs. Proteins such as CELSR2 and FGF5, emphasized in the SMR analysis, displayed strong evidence of colocalization across three identified signature pairs, underscoring a shared pathogenic pathway across these conditions.

## Discussion

In this study, a comprehensive genome-wide pleiotropy analysis was carried out to investigate the genetic correlation and overlap between eGFR and CVDs, uncovering a complex shared genetic foundation. Our detailed investigations identified key pleiotropic loci and genes, including *L3MBTL3*, *MMP24*, and *ABO*, and illuminated crucial biological pathways such as stem cell population maintenance and glutathione metabolism. Additionally, using MR analysis, we indicated a causal relationship between eGFR and VTE, as well as between AF and eGFR. Collectively, the results provide a better understanding of the shared genetic etiology and biological mechanisms that underlie the relationship between eGFR and CVDs, opening new avenues for targeted therapeutic strategies and further research.

Our analysis probed the shared genetic architectures of eGFR and six major CVDs using complementary genetic tools—LDSC, MiXeR, and LAVA—to reveal extensive shared genetic bases across all trait pairs. LDSC highlighted significant negative global *r_g_* between eGFR and VTE and Stroke, contrasting with previous studies that reported no significant associations. This underscores the advantage of our approach and the benefits of a larger sample size. Despite the absence of significant global correlations for certain traits, MiXeR identified substantial overlap for CAD, HF, and AF with eGFR, indicating mixed effect directions even when global correlations are not evident.

LAVA provided further insights into local-*r_g_s*, revealing mixed effect directions not apparent from genome-wide estimates, such as those observed between eGFR and CAD, which exhibited both positively and negatively correlated loci. This suggests a more nuanced genetic relationship than previously recognized. Moreover, employing bivariate MiXeR revealed extensive genetic overlap between eGFR and CAD, supported by LAVA’s local-*r_g_s*, highlighting concordant and discordant effect directions. Our comprehensive analysis confirms the shared genetic foundations between eGFR and various CVDs, indicating multiple common genetic variants influencing critical biological pathways.

Our findings elucidate the shared genetic bases between eGFR and CVDs, attributable to both horizontal pleiotropy and direct causal mechanisms (vertical pleiotropy). Our MR analysis effectively separated potential causal effects from other genetic influences. Notably, the LHC-MR results identified a reverse causal relationship of eGFR on VTE, a finding supported by Yuan et al.^43^ This causal relationship is likely driven by impaired kidney function, which increases coagulation factors and decreases endogenous anticoagulants, collectively promoting thrombus formation. Additionally, our analysis indicates a potential causal influence of AF on eGFR, aligning with previous studies that suggest AF can lead to renal impairments via mechanisms such as thromboembolism and reduced renal microvascular blood flow.^12,44^ However, our analysis did not establish causal relationships for other trait pairs, suggesting that while genetic overlaps between eGFR and CVDs are partially driven by causality, the complexity of these relationships varies across different conditions.

Our cross-trait meta-analysis has identified many genome-wide significant pleiotropic loci associated with eGFR and CVDs. Notably, loci such as 8p23.1 (*BLK*) and 6q25.3 (*SLC22A1*) demonstrated significance across six trait pairs, while others like 12q24.12 (*ALDH2*) and 20q11.22 (*PROCR*) were significant in over 50% of the trait pairs. Specifically, the BLK gene at 8p23.1 enhances insulin synthesis and secretion, linking it to CKD symptoms and complications in nephropathy and diabetes by increasing the production of pro-inflammatory cytokines. BLK is also identified as a susceptibility gene for Kawasaki disease,^45^ a systemic vasculitis that can impact coronary arteries and increase CVDs risks.^46^ Furthermore, the locus *SLC22A1* at 6q25.3 is associated with DN, a severe complication of diabetes characterized by persistent albuminuria, declining eGFR, and elevated arterial blood pressure, which elevates the risk for CVDs and cerebrovascular diseases.^47^ The role of this locus in hypertension underscores its potential as a mediator of underlying disease risks. The consistency of effect alleles in nearly half of the significant SNPs for both eGFR and CVD supports a positive association between these conditions and provides insights into their complex *r_g_*.

Our analysis has identified several key pleiotropic genes in over 50% of the trait pairs, including *L3MBTL3* (6q23.1), *MMP24* (20q11.22), and *ABO* (9q34.2). For example, *L3MBTL3* significantly influences the risk of coronary heart disease (CHD) by modulating adipogenesis, the process of fat cell differentiation. Dysfunctional adipose tissue can lead to ectopic fat accumulation, which promotes insulin resistance and other metabolic disorders, escalating the risk for obesity-related CHD. These metabolic disturbances further amplify the risk of ischemic stroke by intensifying common risk factors (such as hypertension and diabetes). Although *L3MBTL3*’s expression in the kidney is documented, the specific mechanisms by which it affects renal function and structure remain to be fully explored. Matrix metalloproteinase-24 (*MMP24*) plays a key role in extracellular matrix degradation, and is linked with renal tubular atrophy and the severity of alcohol-induced renal injury and fibrosis. Dysregulated *MMP24* expression detrimentally impacts both renal and cardiovascular health. In DN, *MMP24* is implicated in abnormal extracellular matrix accumulation, leading to significant damage to the glomeruli and renal tubules and indirectly impacting eGFR.^48^ Given the close association of DN with T2D, a prominent risk factor for CVDs, *MMP24*’s role in DN could indirectly heighten the risk of CVDs.^49^ The *ABO* gene, vital for determining blood group phenotypes, also affects crucial hemostatic factors such as VWF and factor VIII (FVIII), which are essential for platelet adhesion at injury sites. Individuals with non-O blood types generally exhibit higher VWF levels and are more susceptible to VTE.^50^ In addition, researchers have discovered a new connection between *ABO* and primary glomerulonephritis, especially IgA nephropathy, where certain blood groups are linked to an increased risk of renal function decline.^51^ These findings highlight the complex interaction of genetics with the pathophysiology of renal and CVDs, which provides a strong basis for further study of the mechanisms and risks associated with genetic diseases.

Our study also examines the role of shared genetic determinants in two critical biological pathways, including stem cell population maintenance and glutathione catabolism, and their implications for kidney function and cardiovascular health. In the stem cell maintenance pathway, *MMP24*^52^, *PSRC1*^53^, and *CHMP1A*^54^ play pivotal roles by regulating the stem cell cycle, ensuring cellular quiescence, and preventing premature differentiation, respectively. These molecular actions are crucial for promoting tubular cell growth and enhancing renal function, mainly through the secretion of regenerative factors such as bone morphogenetic protein-7 and hepatocyte growth factor.^55^ The effectiveness of stem cell therapy in heart repair, particularly in those with elevated levels of inflammation, further underscores the importance of robust stem cells for optimal cardiovascular and renal outcomes. In the glutathione pathway, *DPEP1*^56^, *ORMDL3*, *MRPS21*, and *CPS1*^57,58^ are crucial for maintaining cellular redox balance and defending against oxidative stress, and they are vital for stroke prevention and kidney function maintenance.^59,60^ Variations in kidney function, reflected by changes in eGFR, can significantly affect glutathione turnover and its protective roles, directly linking renal and cardiovascular health.^61^ Our findings suggest that targeted modulation of these key genes could significantly enhance kidney and heart repair and regeneration, thus improving eGFR and decreasing the risk of CVDs.

Our study has some limitations. Firstly, the variation in sample sizes across disorders, from 511,634 cases for PAD to 1,500,861 for VTE, restricted our ability to detect pleiotropic effects in less-represented diseases. Future studies with larger, more balanced samples will likely improve our capacity to discern these effects more precisely. Additionally, including subjects with concurrent eGFR abnormalities and CVDs may have introduced bias in our assessment of genetic overlap between these conditions. Our analysis focused on shared genetic variants, excluding rare mutations that could further elucidate the complex genetic relationships involved. Moreover, the concentration on individuals of European ancestry limits the generalizability of our findings to other populations. It is crucial to conduct cross-ancestry follow-up studies to determine whether these results hold across diverse ethnic groups. Overcoming these limitations will require enhancing the diversity of GWAS populations and conducting more extensive experimental validations and cohort analyses.

## Conclusion

Together, this study has significantly improved our understanding of the genetic basis for the role of eGFR in susceptibility to major CVDs. By identifying crucial genetic components, including key pleiotropic loci and genes such as *L3MBTL3*, *MMP24*, and *ABO*, we elucidate potential shared mechanisms that may influence critical biological pathways associated with CVDs, notably stem cell population maintenance and glutathione metabolism. These results provide a better understanding of the genetic basis of CVDs and provide a basis for the development of more targeted therapeutic and preventive strategies that could potentially transform cardiovascular care.

## Disclosure

All authors declare no competing interests.

## Data Statement

The study used only openly available GWAS summary statistics on estimated glomerular filtration rate and six major cardiovascular diseases that have originally been conducted using human data. GWAS summary statistics on eGFR are available at https://www.uni-regensburg.de/medizin/epidemiologie-praeventivmedizin/genetische-epidemiologie/gwas-summary-statistics/index.html. GWAS summary statistics on AF, HF, and Stroke are available at the GWAS Catalog (GCST90104539, GCST009541, and GCST90104539). GWAS summary statistics on CAD and PAD are publicly available for download at the Cardiovascular Disease Knowledge Portal (CVDKP) website: https://cvd.hugeamp.org/datasets.html. GWAS summary statistics on VTE are obtained from the deCODE genetics website: https://www.decode.com/ summarydata/. Blood-based cis-pQTL from UKB-PPP are obtained from https://www.synapse.org/Synapse:syn51365303.

## Data Availability

The study used only openly available GWAS summary statistics on estimated glomerular filtration rate and six major cardiovascular diseases that have originally been conducted using human data. GWAS summary statistics on eGFR are available at https://www.uni-regensburg.de/medizin/epidemiologie-praeventivmedizin/genetische-epidemiologie/gwas-summary-statistics/index.html. GWAS summary statistics on AF, HF, and Stroke are available at the GWAS Catalog (GCST90104539, GCST009541, and GCST90104539). GWAS summary statistics on CAD and PAD are publicly available for download at the Cardiovascular Disease Knowledge Portal (CVDKP) website: https://cvd.hugeamp.org/datasets.html. GWAS summary statistics on VTE are obtained from the deCODE genetics website: https://www.decode.com/summarydata/. Blood-based cis-pQTL from UKB-PPP are obtained from https://www.synapse.org/Synapse:syn51365303.

## Acknowledgements

This study was supported by the Natural Science Foundation of China Excellent Young Scientists Fund (Overseas) (Grant no. K241141101), Guangdong Basic and Applied Basic Research Foundation for Distinguished Young Scholars (Grant no. 24050000763), Shenzhen Pengcheng Peacock Plan, Shenzhen Basic Research General Projects of Shenzhen Science and Technology Innovation Commission (Grant no. JCYJ20230807093514029) (To Y.F.), National Natural Science Foundation of China (Grant no. 82260073); Tianshan Talent Cultivation Program Project of Xinjiang Uygur Autonomous Region (Grant no. 2022TSYCLJ0028) (To Y.Y.), and Center for Computational Science and Engineering at Southern University of Science and Technology. The funder had no role in the design, implementation, analysis, interpretation of the data, approval of the manuscript, and decision to submit the manuscript for publication.

## Author contributions

J.Q., Y. F., Y.Y., J.X., and S.P. conceptualized and supervised this project and wrote the manuscript. J.Q., K.Y., and Y.Y. performed the main analyses and wrote the manuscript. J.Q., X.Y., Y.Y., L.Z., and Y.L. performed the statistical analysis and assisted with interpreting the results. M.C., W.X., and Y.Y. provided expertise in cardiovascular biology and GWAS summary statistics. All authors provided intellectual content and approved the final version of the manuscript.

