## Supplementary file for "Disentangling shared genetic etiologies for kidney function and cardiovascular diseases"

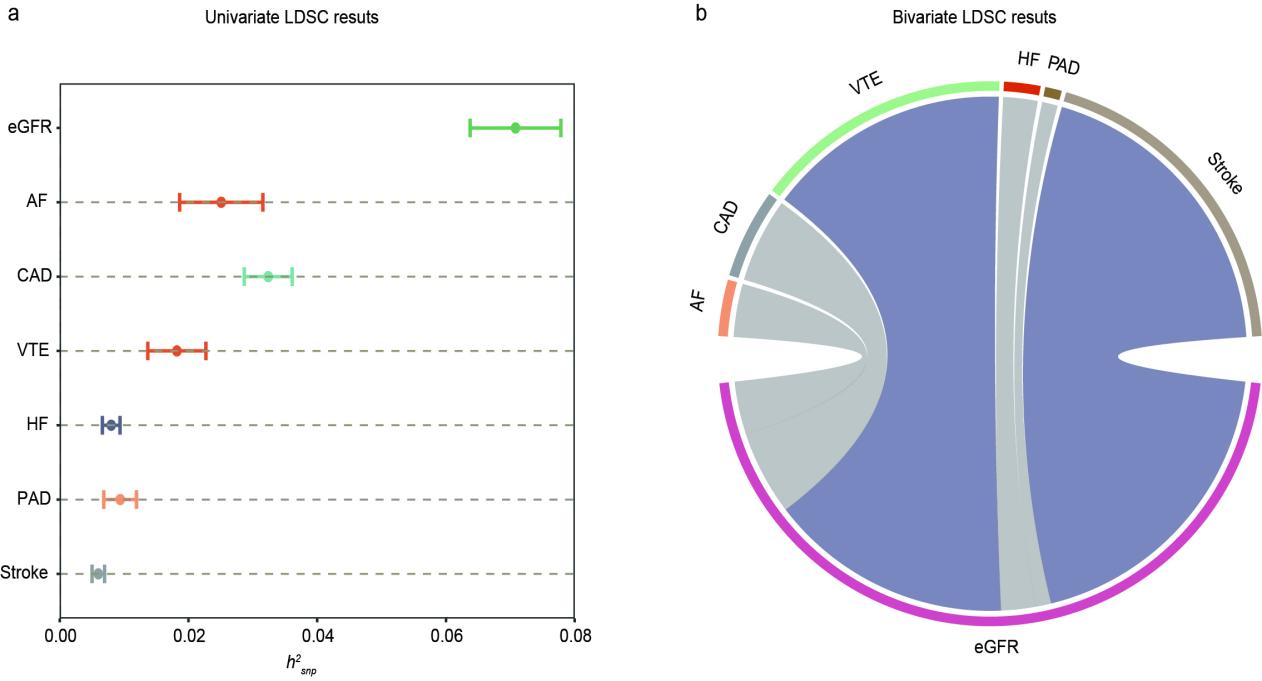

**Supplementary Figure 1: Supplemental LDSC figures for estimated glomerular filtration rate and six major cardiovascular diseases.**

(a) Error-bar plot of the SNP-based heritability (*h^2^_SNP_*) point estimates for eGFR and CVDs, computed using univariate LDSC. (b) Network visualization of the Bonferroni-corrected significant global genetic correlations (*r_g_*) between eGFR and CVDs, computed using bivariate LDSC.

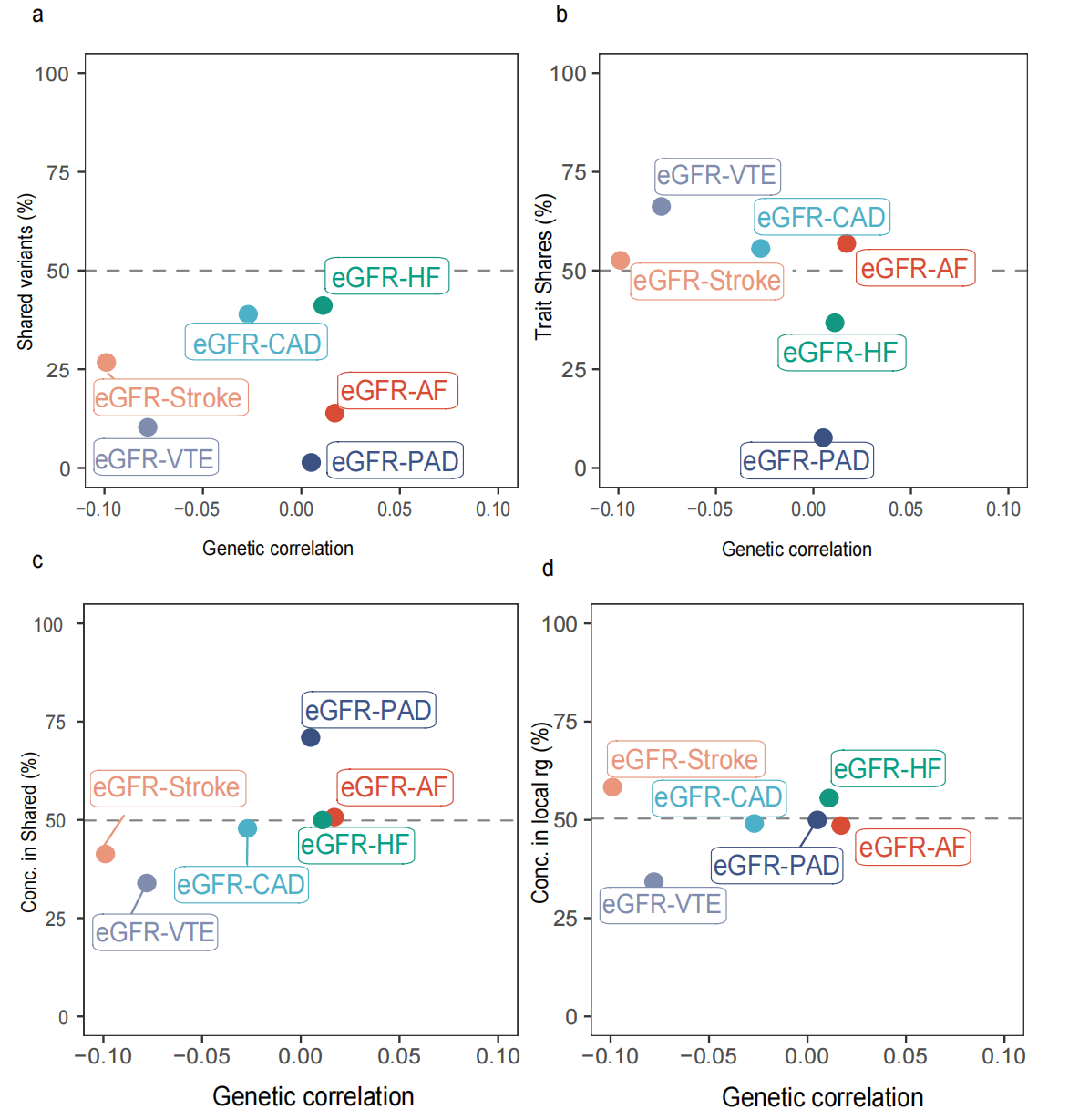

**Supplementary Figure 2: Genetic correlation and genetic overlap between estimated glomerular filtration rate and six major cardiovascular diseases**

a: Genetic correlation estimated by LDSC and percentage of eGFR variants shared with CVDs estimated by MiXeR. b: Percentage of CVDs variants shared with eGFR. c: Percentage of CVD variants with consistent direction of effect shared with eGFR. d: Percentage of local genetic correlation from LAVA with consistent direction of effect on the y-axis. eGFR, estimated glomerular filtration rate; AF, atrial fibrillation; CAD, coronary artery disease; VTE, venous thromboembolism; HF, heart failure; PAD, peripheral arterial disease.

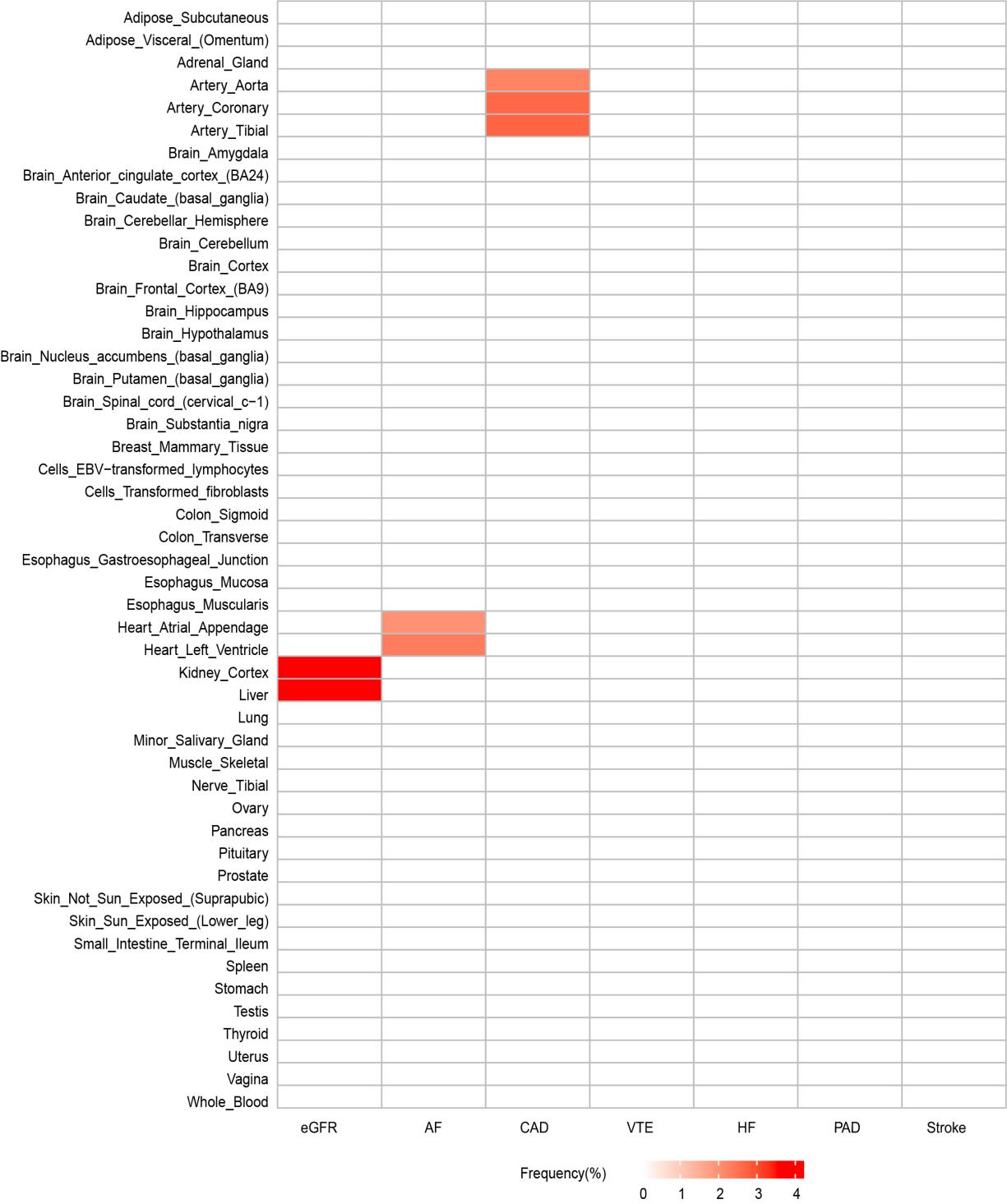

**Supplementary Figure 3: Results of multi-tissue analysis of estimated glomerular filtration rate and six cardiovascular diseases using gene expression data.**

Tissue type-specific enrichment of single nucleotide polymorphism (SNP) heritability for eGFR and CVDs in 49 tissues in GTEx v8 estimated using stratified LDSC applied to specifically expressed genes (LDSC-SEG). Enrichment was measured by significance test in a one-sided Z test and is shown as -log10 (*P*). Multiple testing was performed on the tissues tested, and only tissues with significant association (FDR < 0.05) were shown.

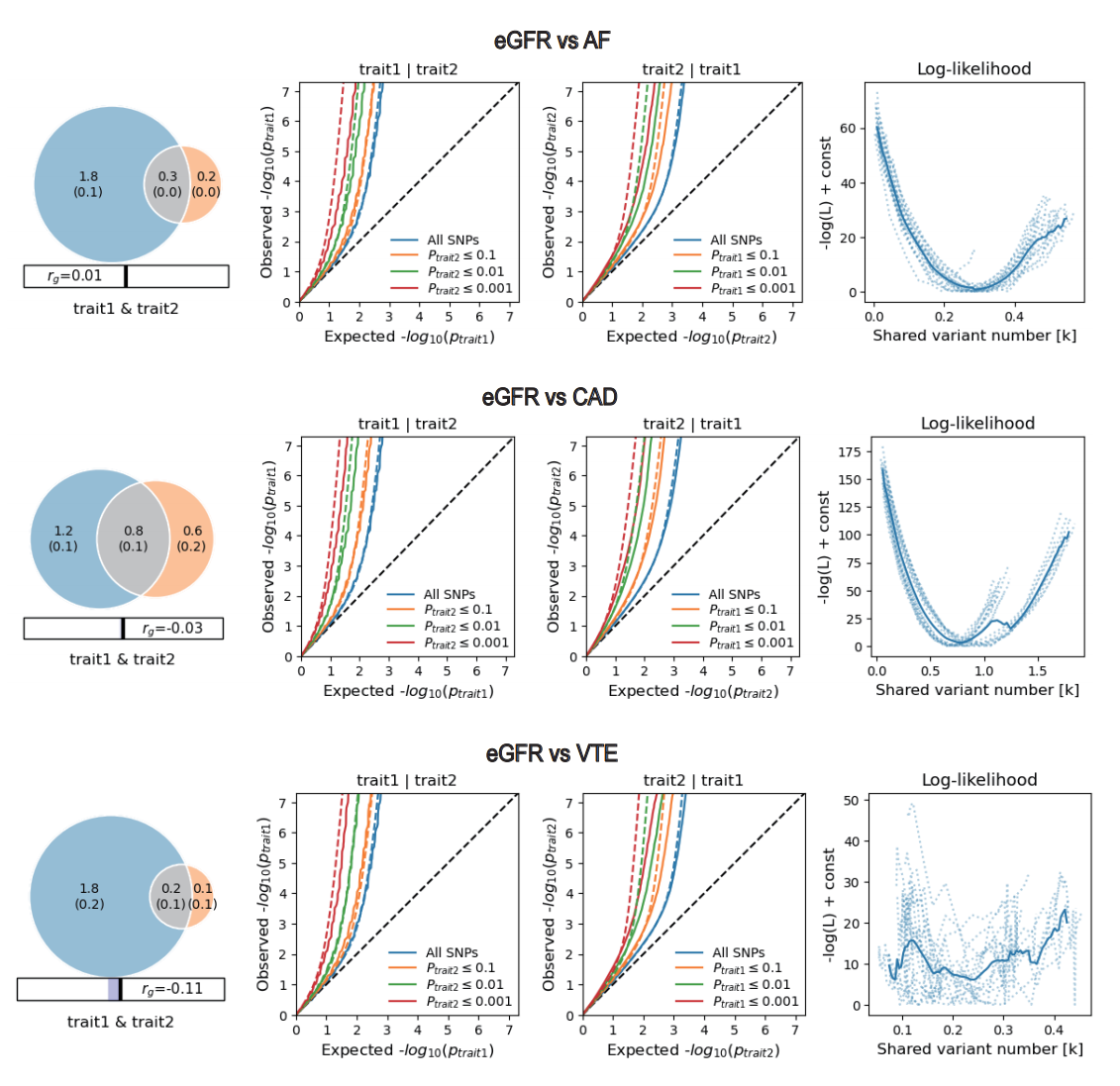

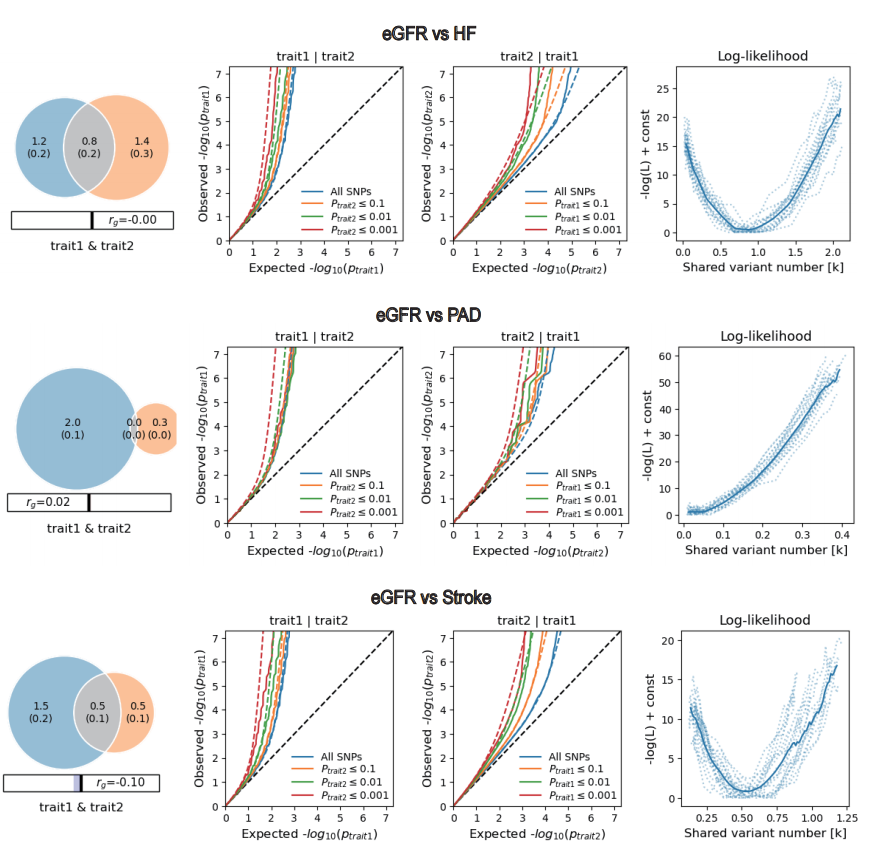

**Supplementary Figure 4: Supplemental MiXeR figures for each of estimated glomerular filtration rate and six major cardiovascular diseases.**

Left: The estimated number of shared causal variants between eGFR and CVDs calculated by MiXeR is shown in gray, and rg is the genetic correlation estimated by LDSC. Middle: The conditional Q-Q plot shows the relationship between the expected (x-axis) and observed (y-axis) significance of variants in the original phenotype when the variants are stratified by the p-value in the conditional phenotype. It shows the nominal and empirical −log10-transformed *P*-values in the primary phenotype as a function of the significance of association of SNPs in the secondary phenotype at levels of *P* ≤ 1.00 (all SNPs, blue line), *P* ≤ 0.1 (red line), *P* ≤ 0.01 (yellow line), and *P* ≤ 0.001 (purple line). The dashed lines show the model predictions for each stratum. The black dashed line is the expected Q-Q plot under the null hypothesis. The increased leftward deflection from the no association line for strata with higher significance in the conditional phenotype indicates the presence of polygenic overlap. Middle left: CVDs conditional on eGFR. Middle right: eGFR conditional on CVDs. Points on the Q-Q plot are weighted according to the LD structure, randomly pruned using n=64 iterations, and an LD threshold of r^2^ = 0.1. Right: The log-likelihood curve highlights the model fit by plotting the negative log-likelihood function (lower values indicate better model fit) versus the π12 parameter (the amount of contributed variance shared between the two traits). The remaining model parameters were constrained to their fitted values. π12 on the log-likelihood plot ranges from the smallest possible value π12 = rg*sqrt(π1u, π2u) that is compatible with the estimated genetic correlation to the largest possible value π12 = min(π1u, π2u) that corresponds to the minimum total polygenicity between the two traits. The minimum point represents the best-fitting model estimate of the amount of contributed variance shared between the two traits.

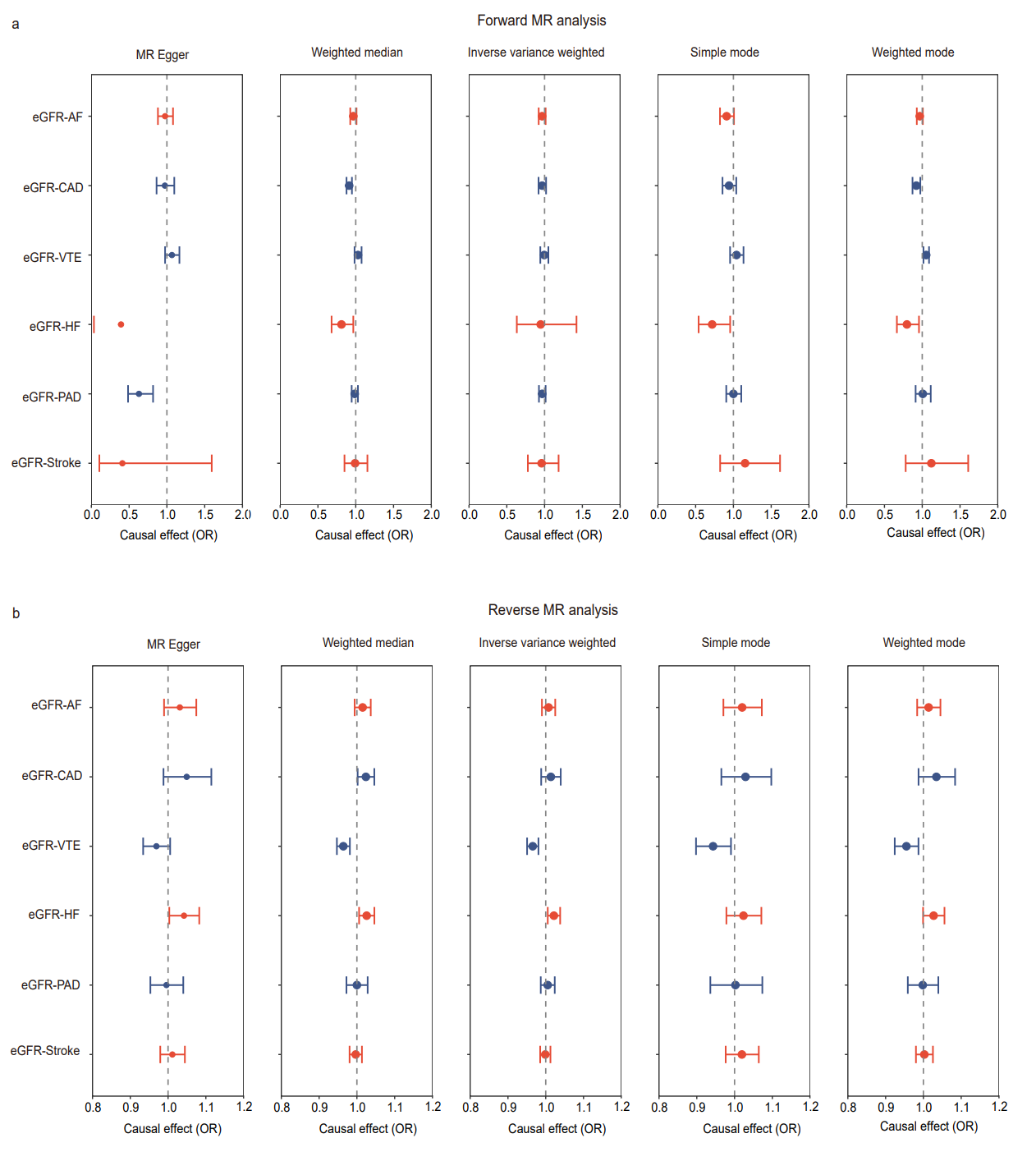

**Supplementary Figure 5: Forest plots of bidirectional causal relationships between estimated glomerular filtration rate and six major cardiovascular diseases.**

Analyses were performed using fixed- and multiplicative random-effects IVW, MR-Egger, weighted median, simple models, and weighted model methods. (a) Forward MR analysis: causal effect of eGFR on CVDs. (b) Reverse MR analysis: causal effect of CVDs on eGFR. Estimates and 95% confidence intervals are presented as square points and error bars, respectively. On the x-axis, the odds ratio (OR) is the effect of a one standard deviation increase in exposure on the outcome, and the vertical dashed line represents the reference at OR = 1. P value less than 0.05 were considered significant. eGFR, estimated glomerular filtration rate; AF, atrial fibrillation; CAD, coronary artery disease; VTE, venous thromboembolism; HF, heart failure; PAD, peripheral arterial disease; CI, confidence interval; IVW, inverse variance weighting; OR, odds ratio.

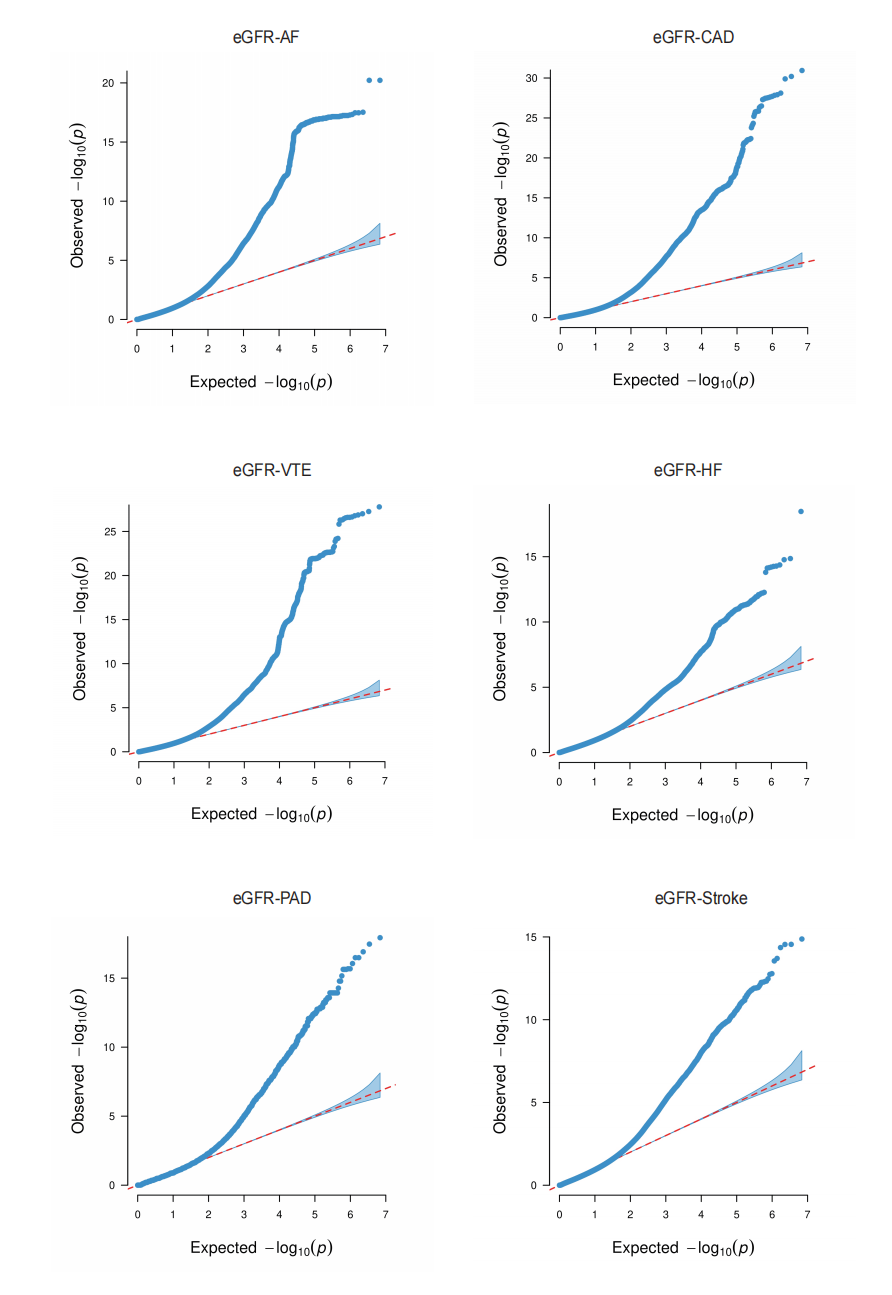
**Supplementary Figure 6: Quantile-quantile (Q-Q) plots for PLACO results of estimated glomerular filtration rate and six major cardiovascular diseases.**

Q-Q plots depicts expected -log10 *P*-values (x-axis) against observed -log10 *P_PLACO_*-values (y-axis). Red dots indicate significant pleiotropic variants (*P_PLACO_* < 5×10^-8^). eGFR, estimated glomerular filtration rate; AF, atrial fibrillation; CAD, coronary artery disease; VTE, venous thromboembolism; HF, heart failure; PAD, peripheral arterial disease.

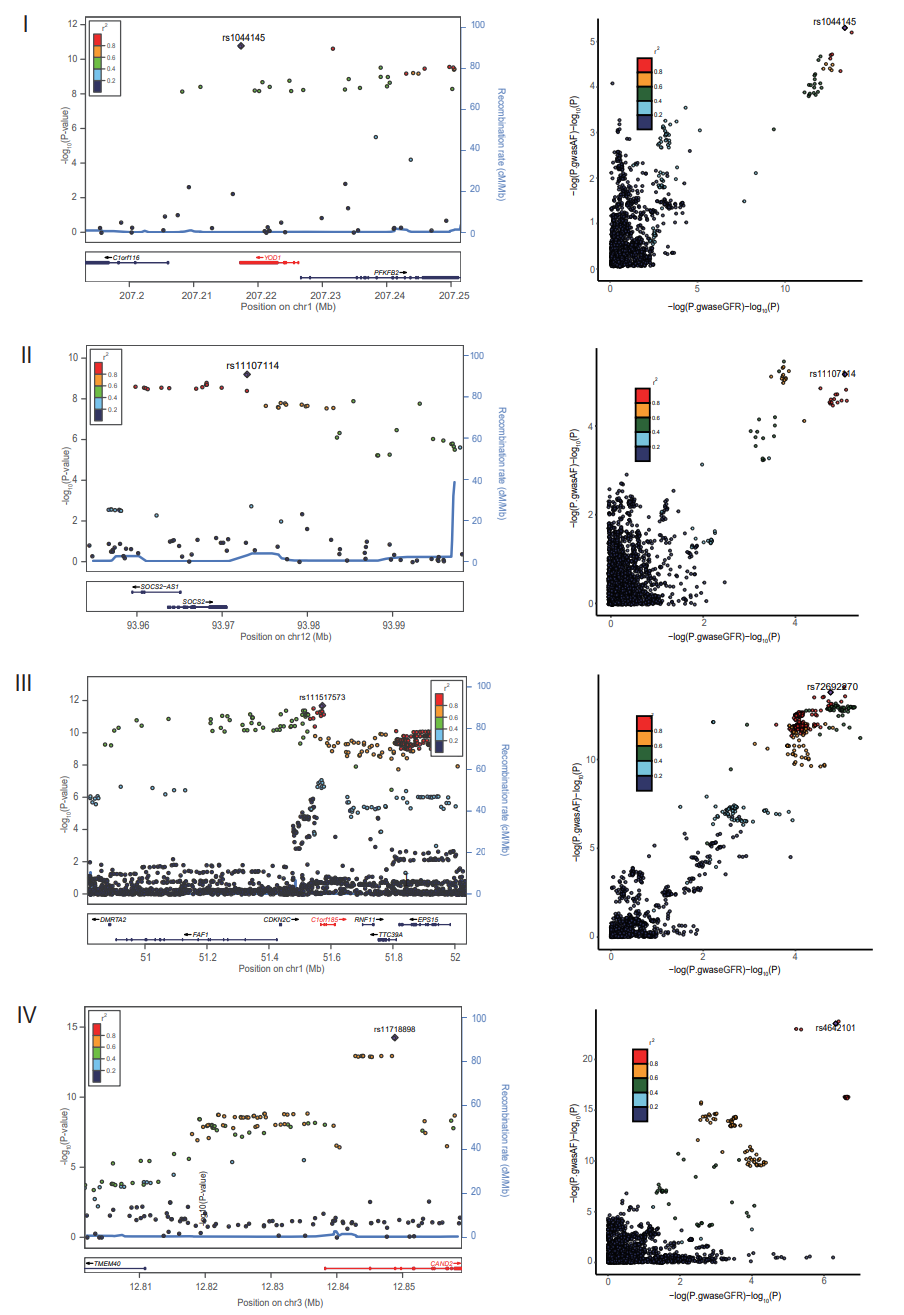

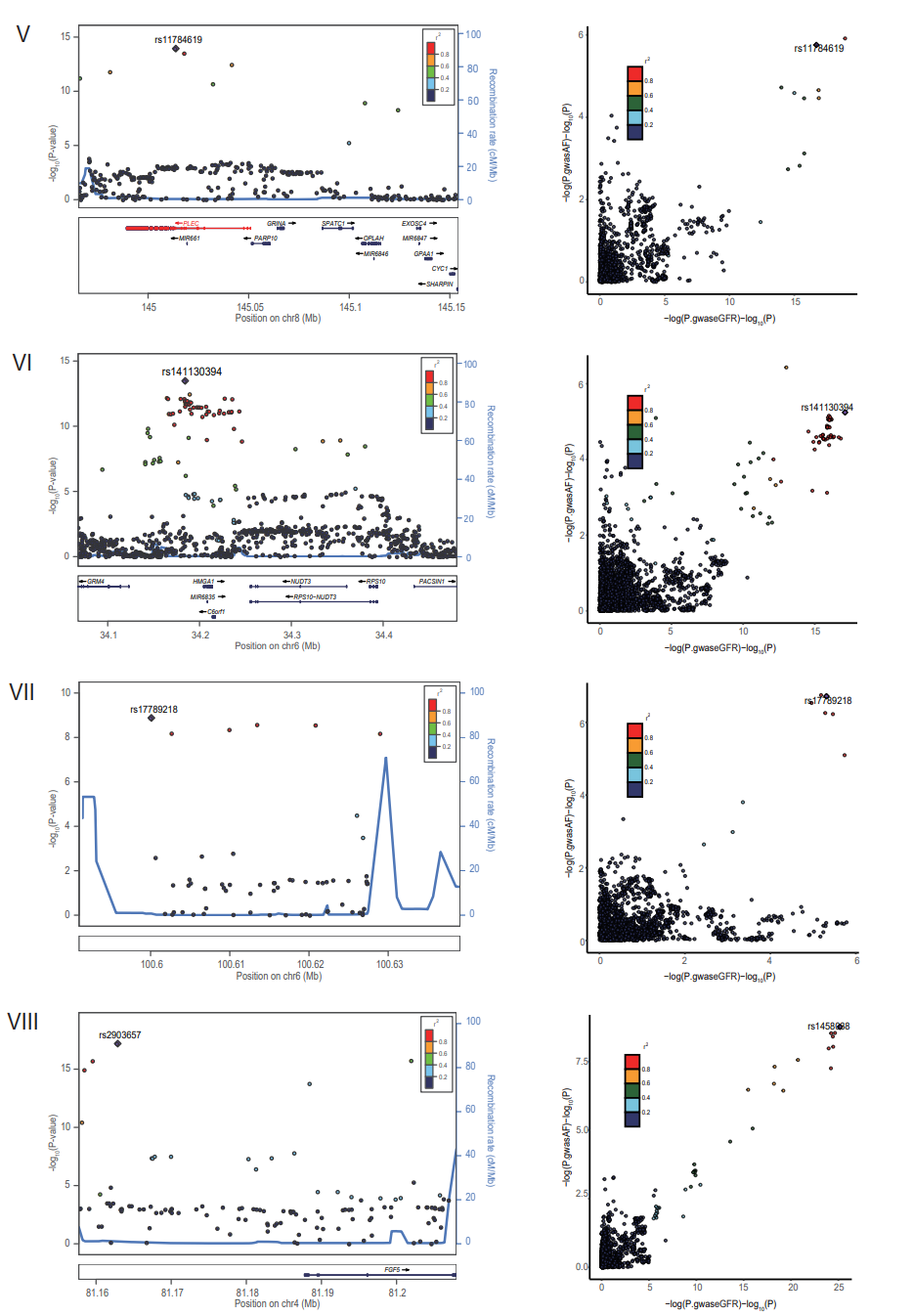

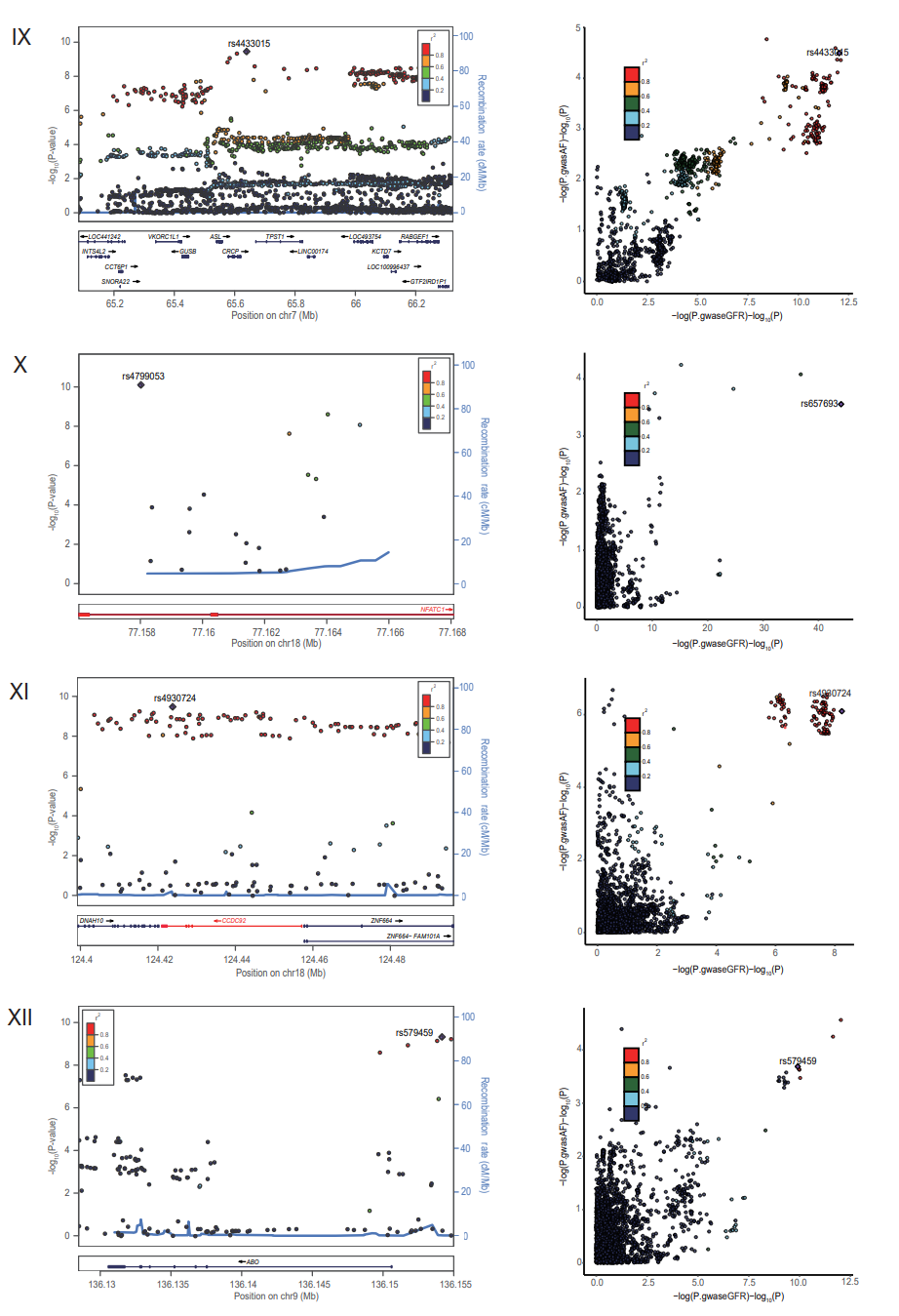

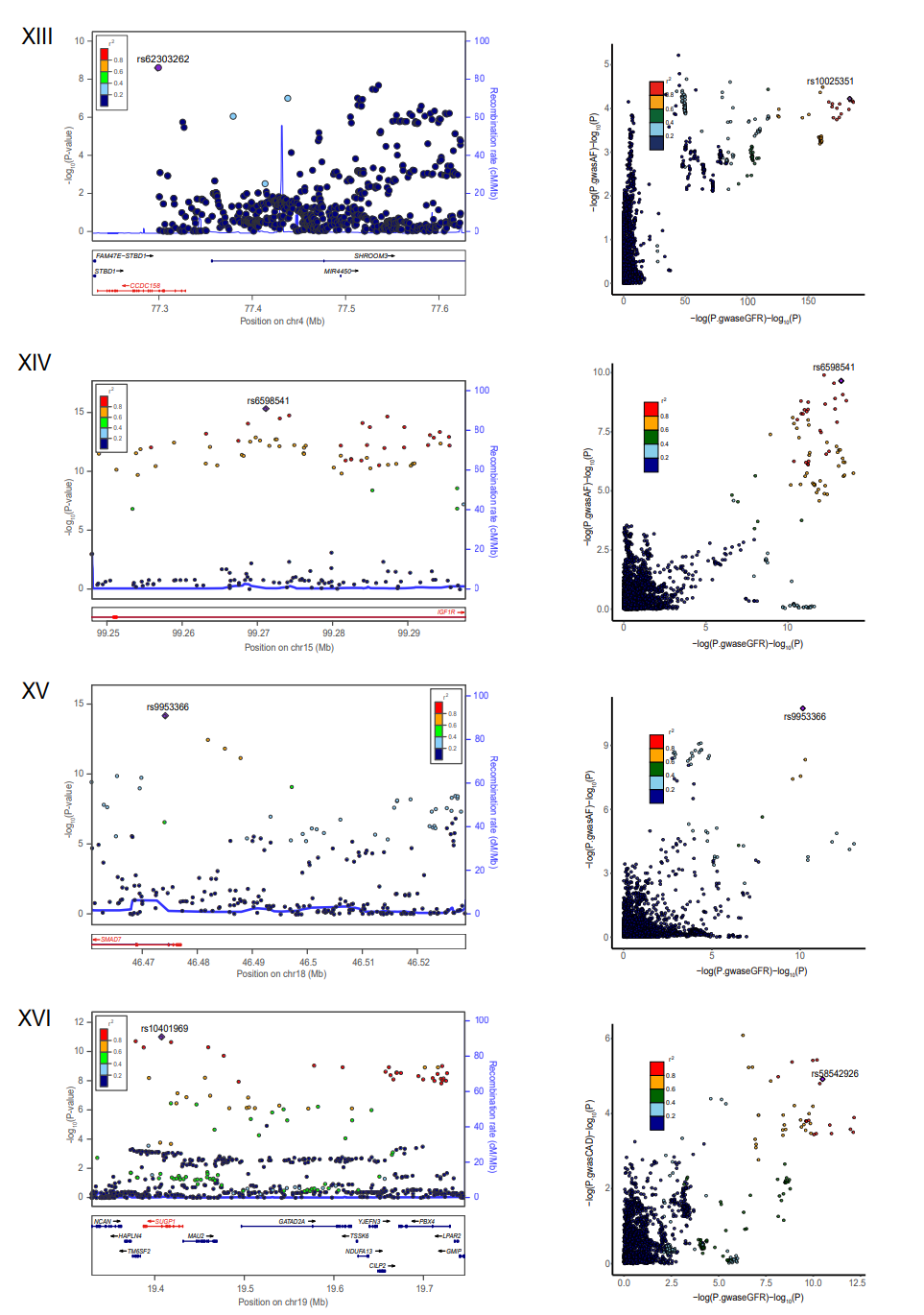

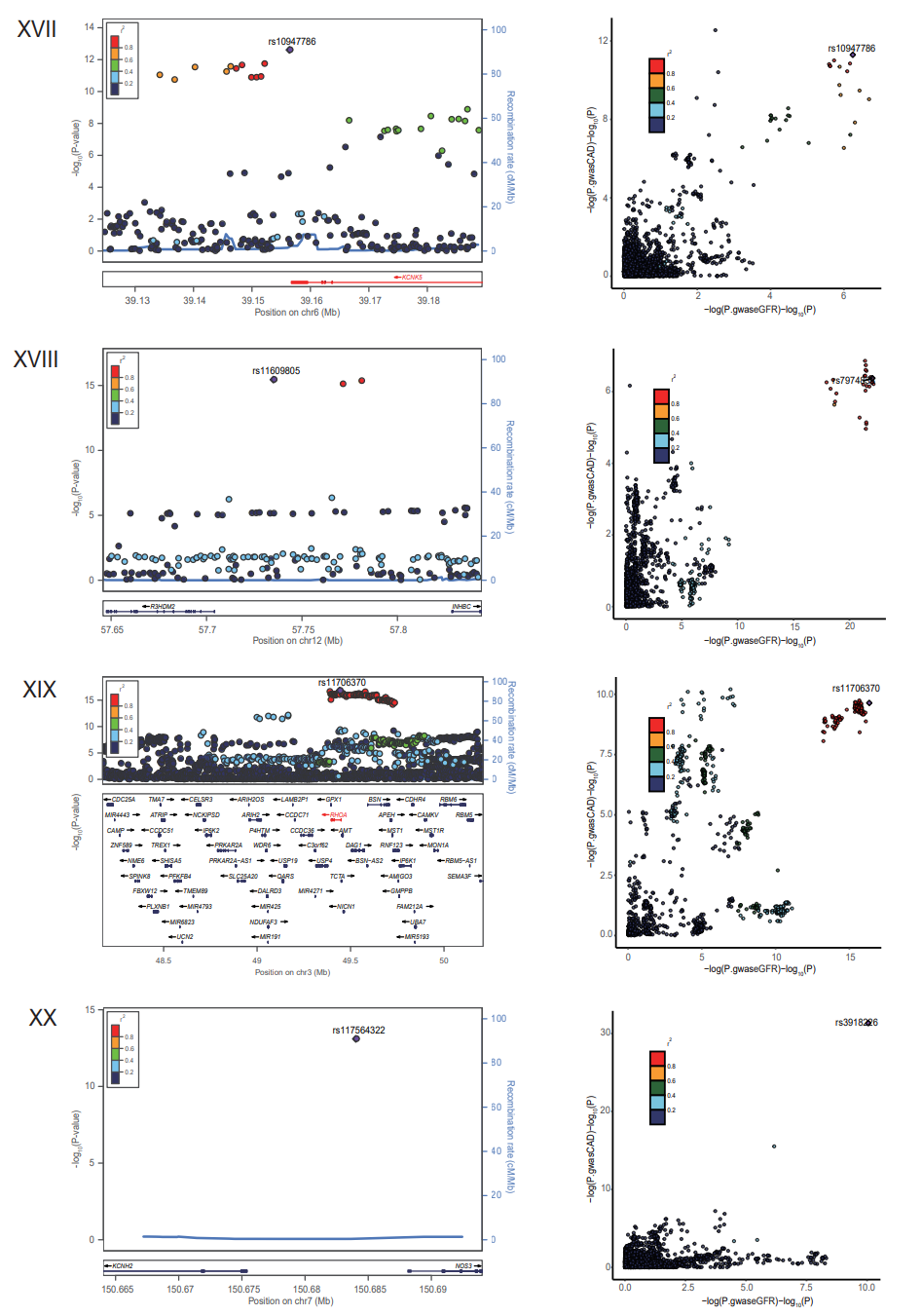

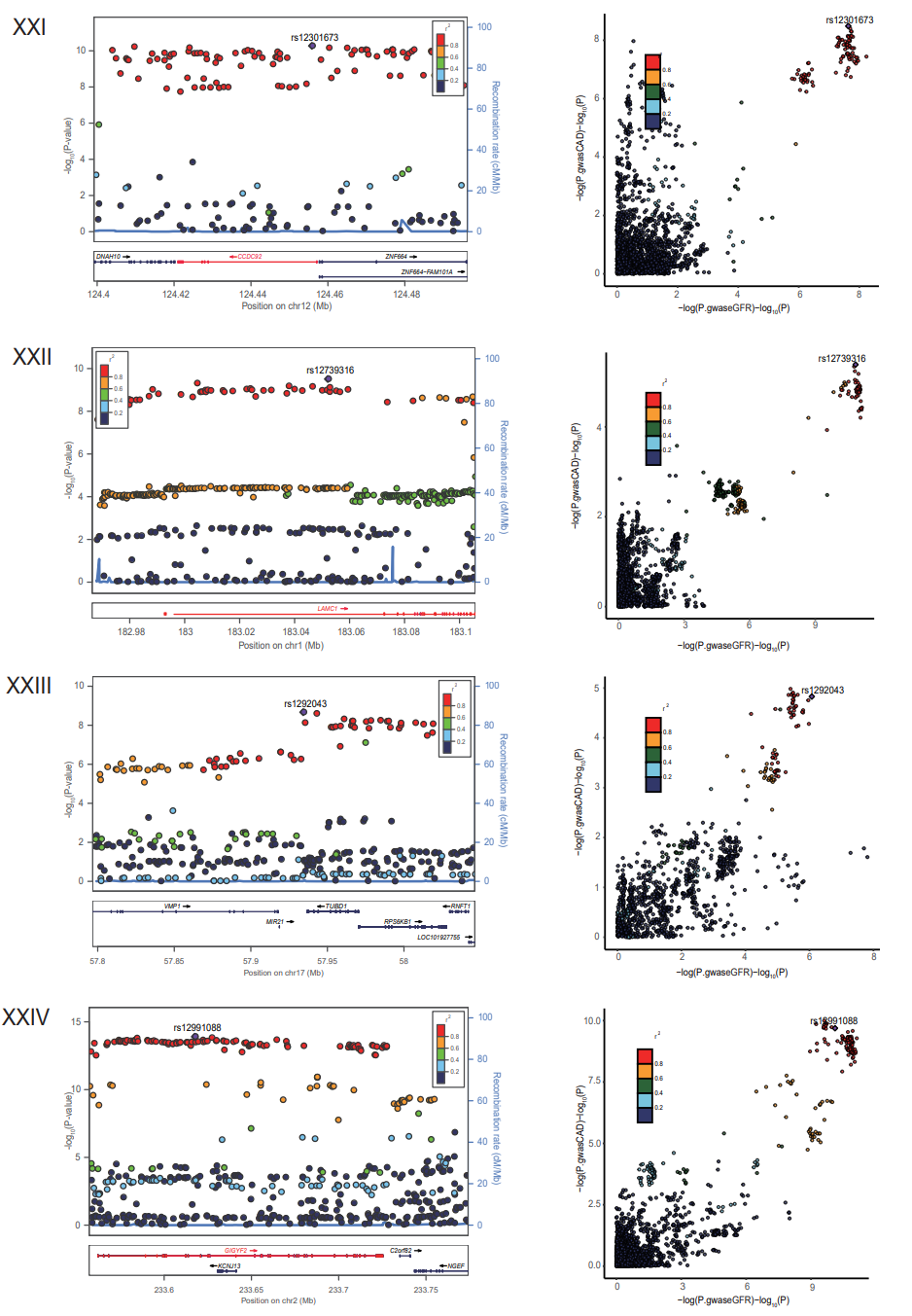

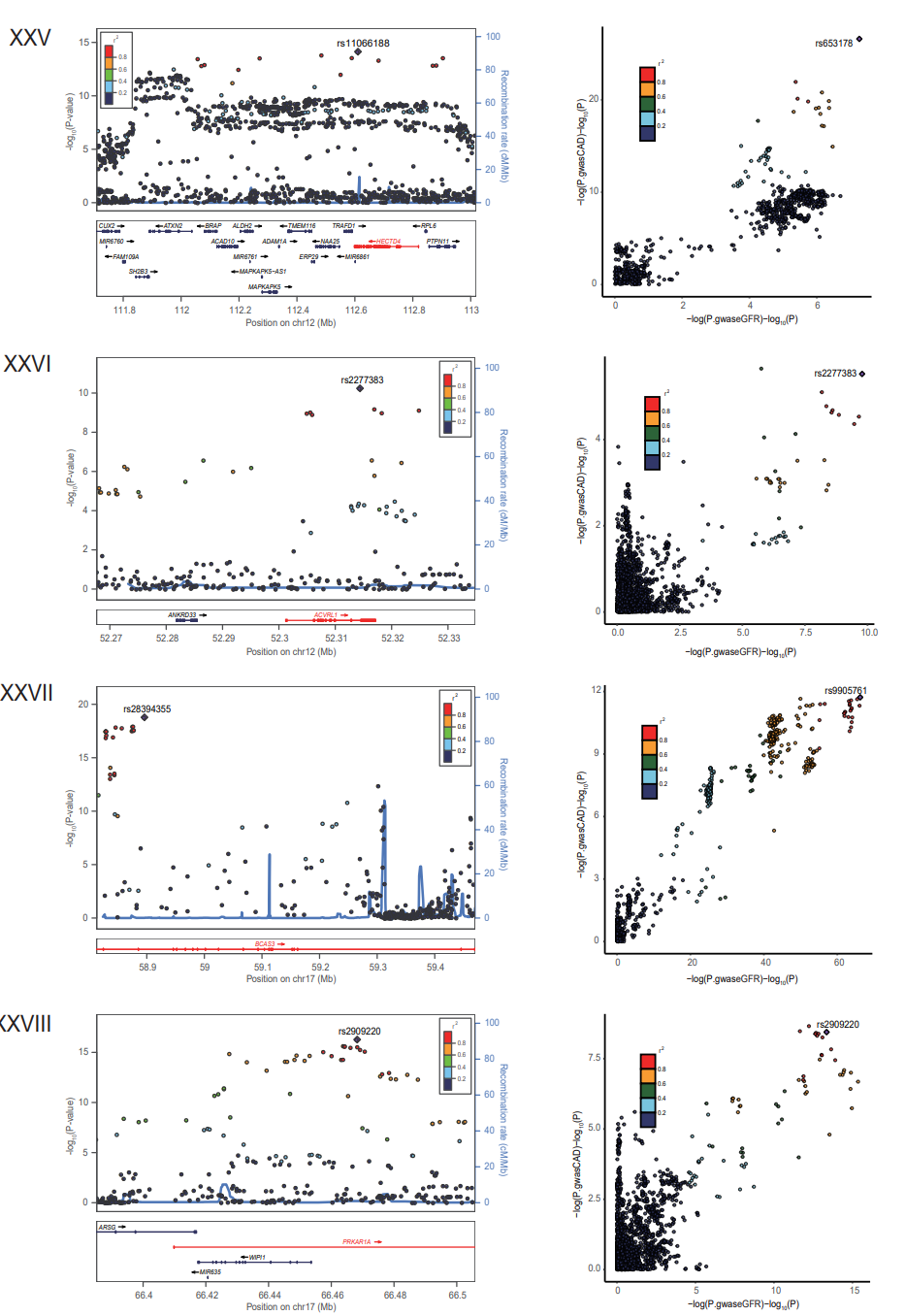

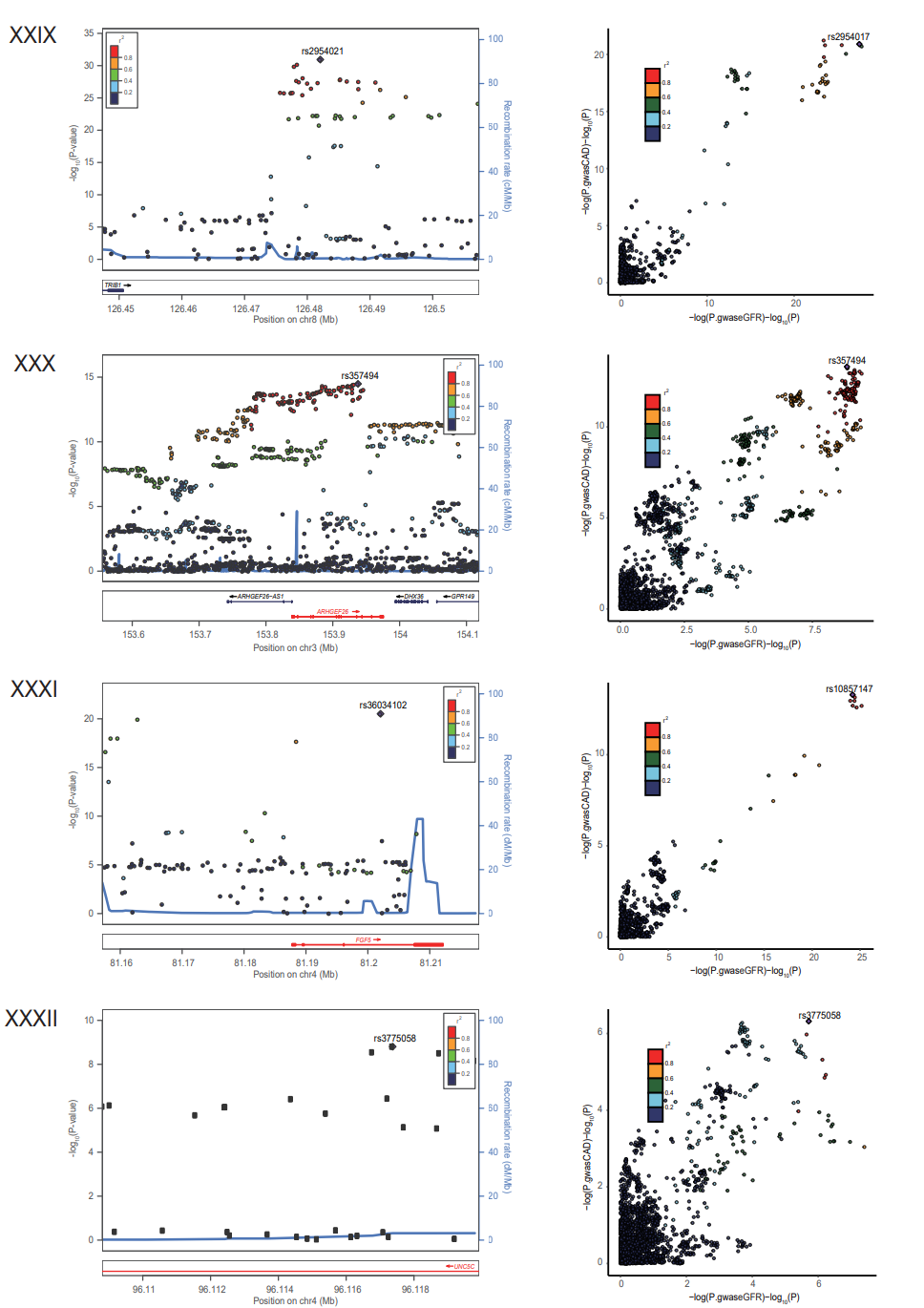

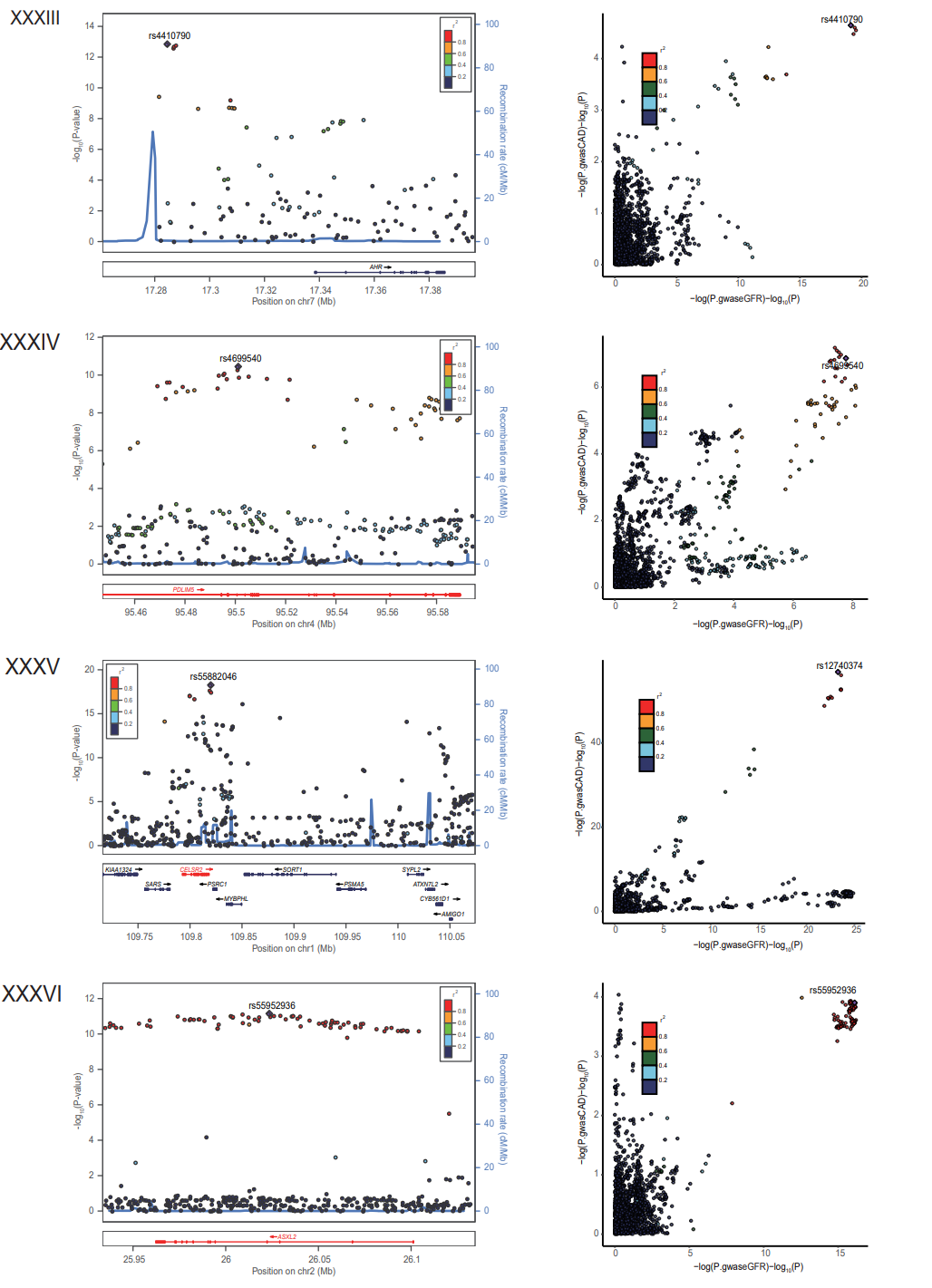

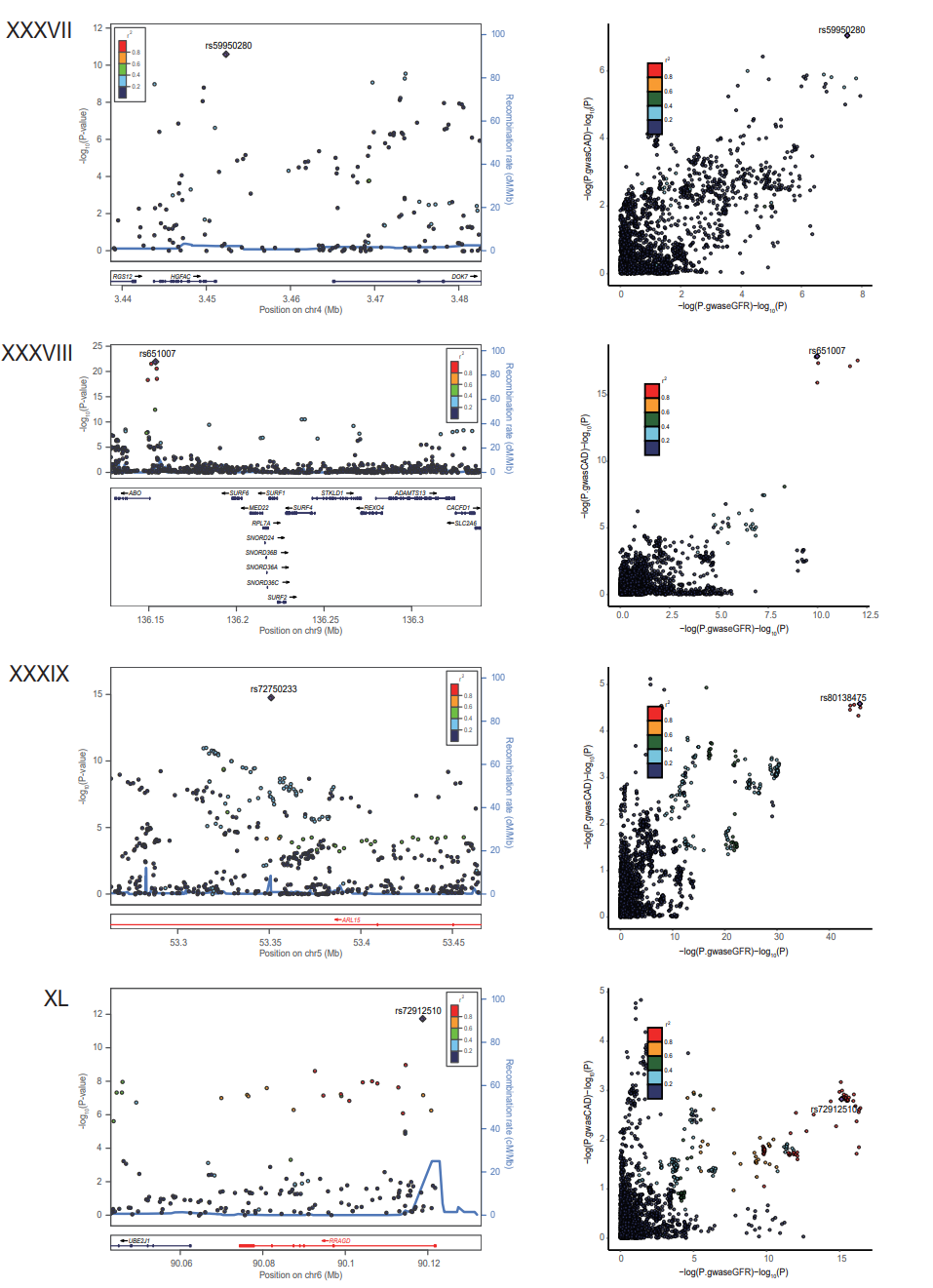

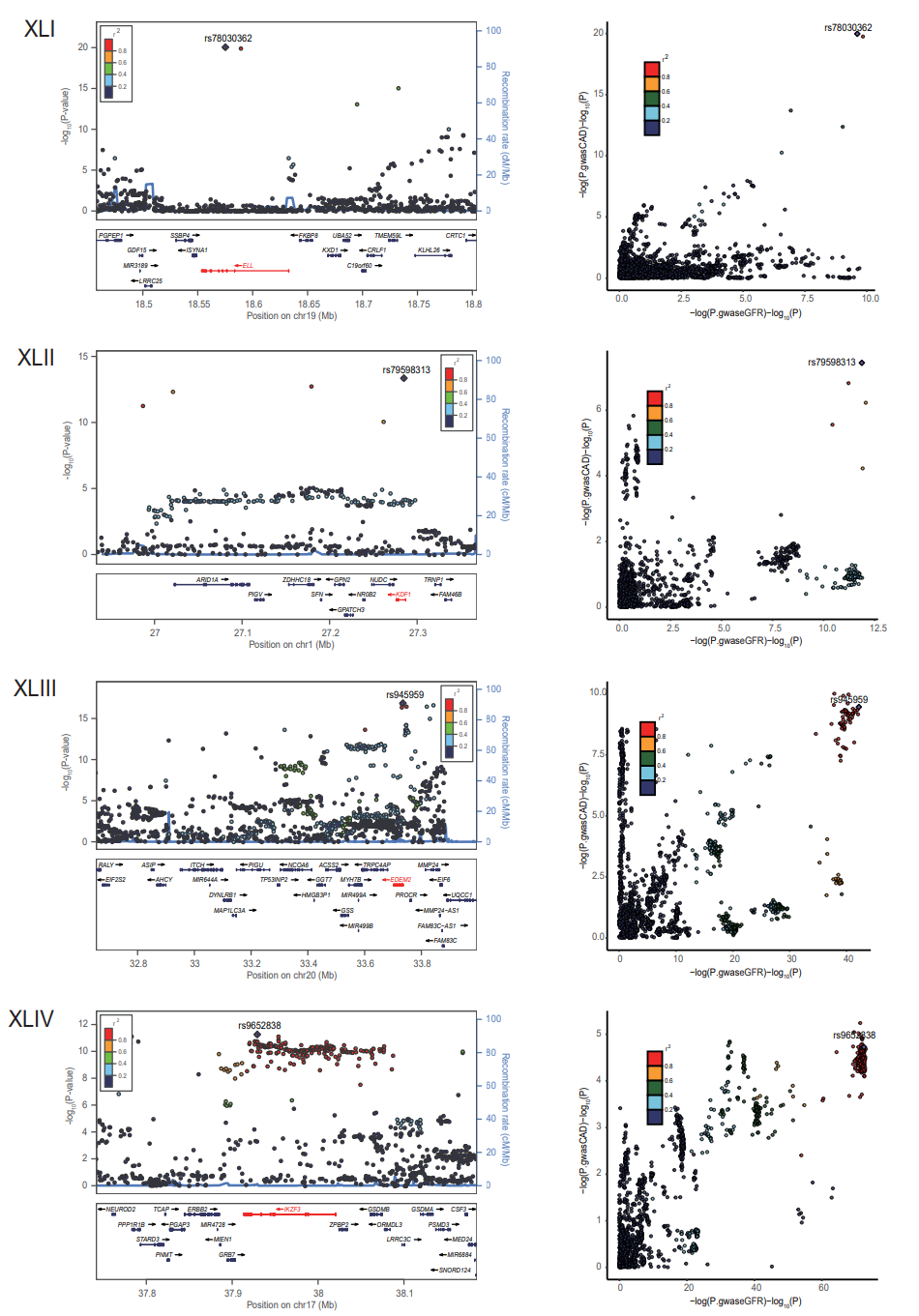

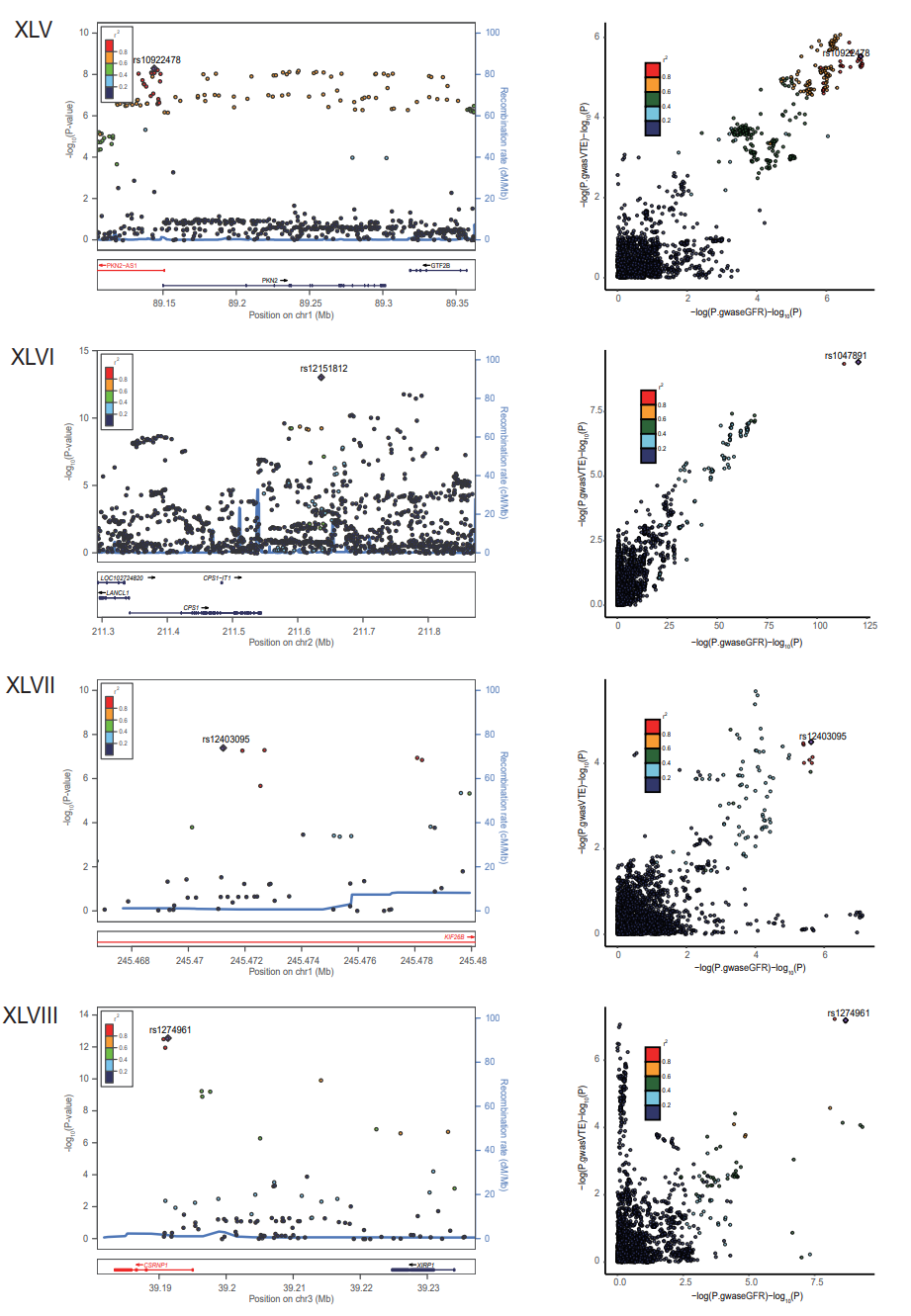

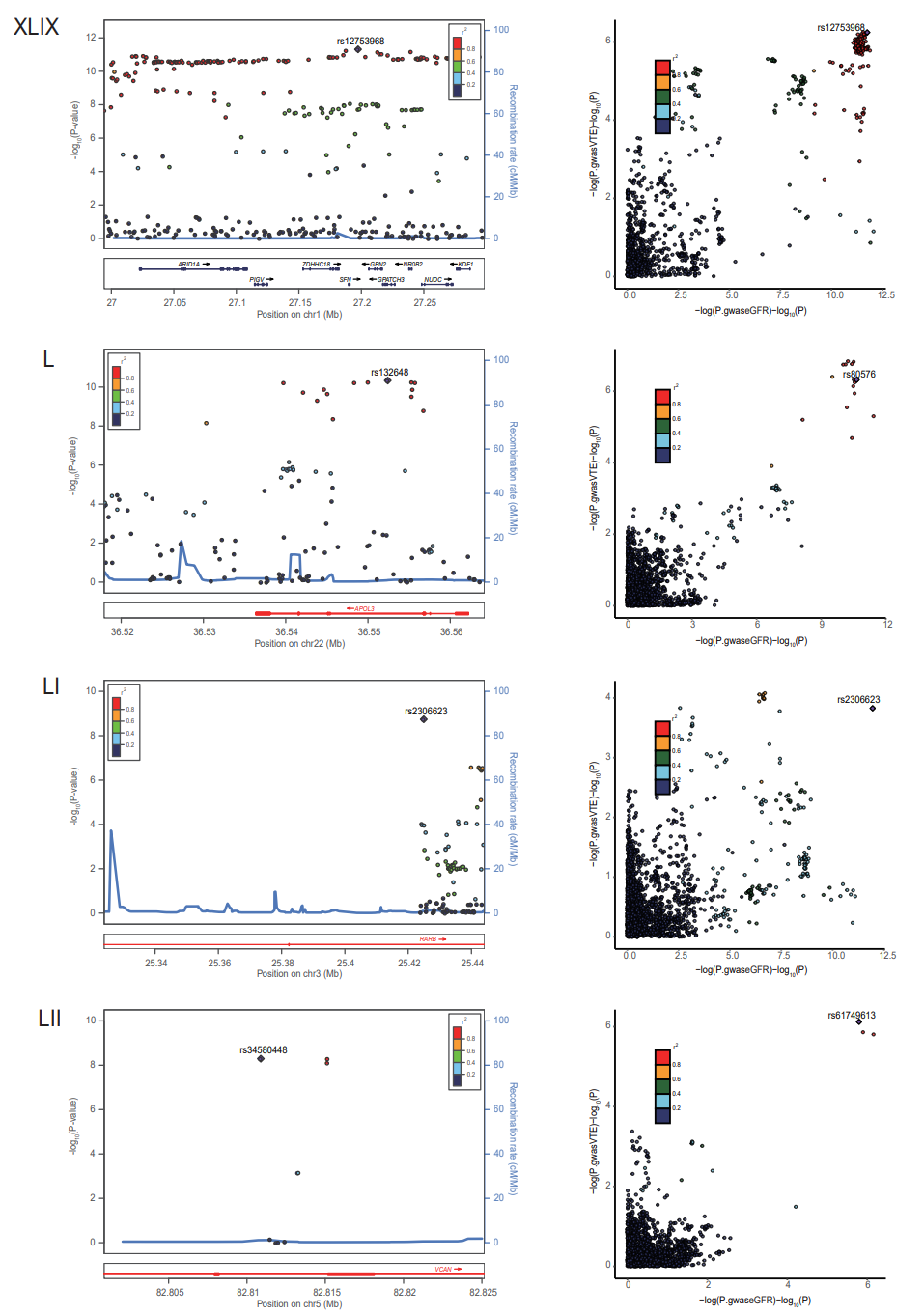

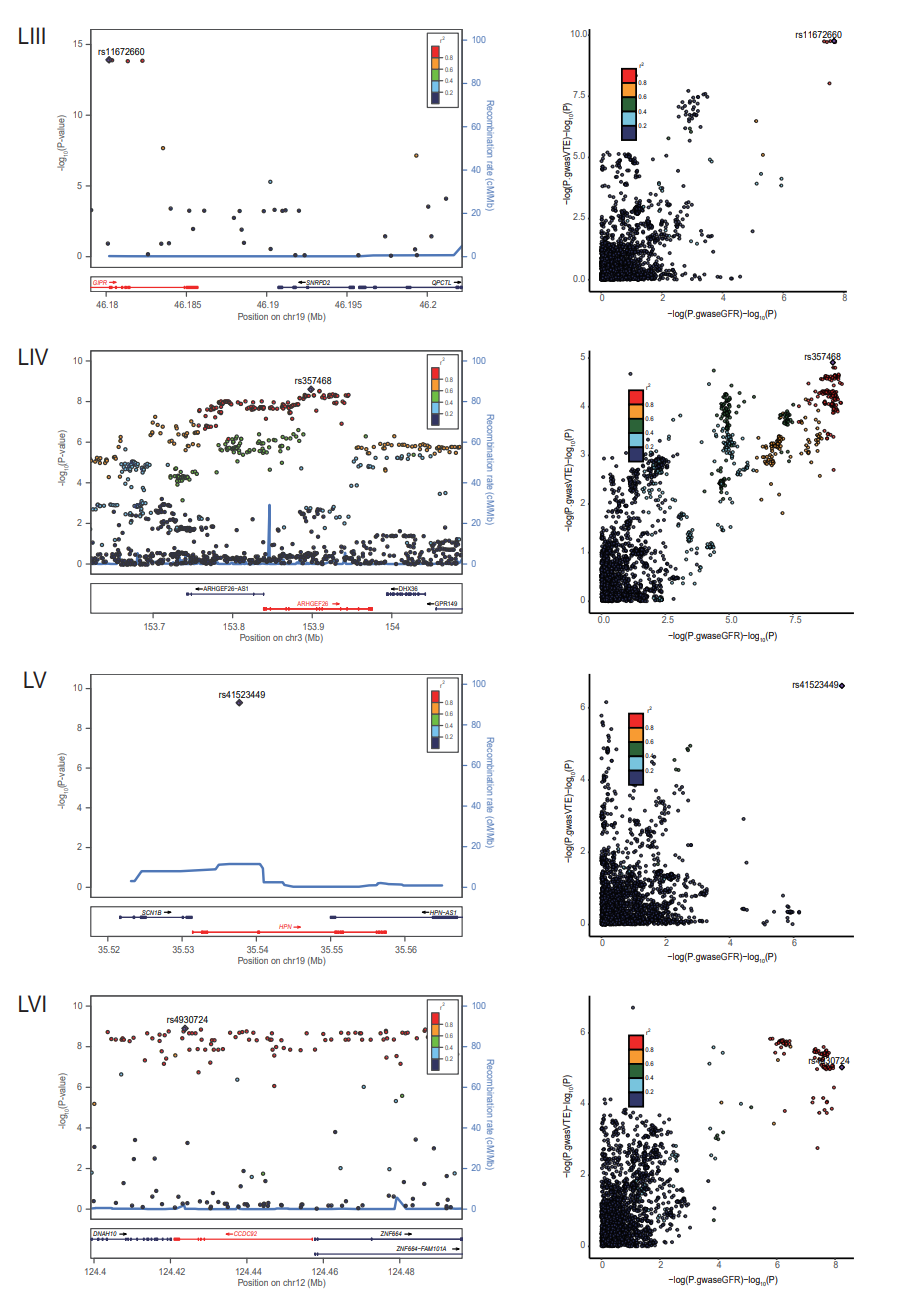

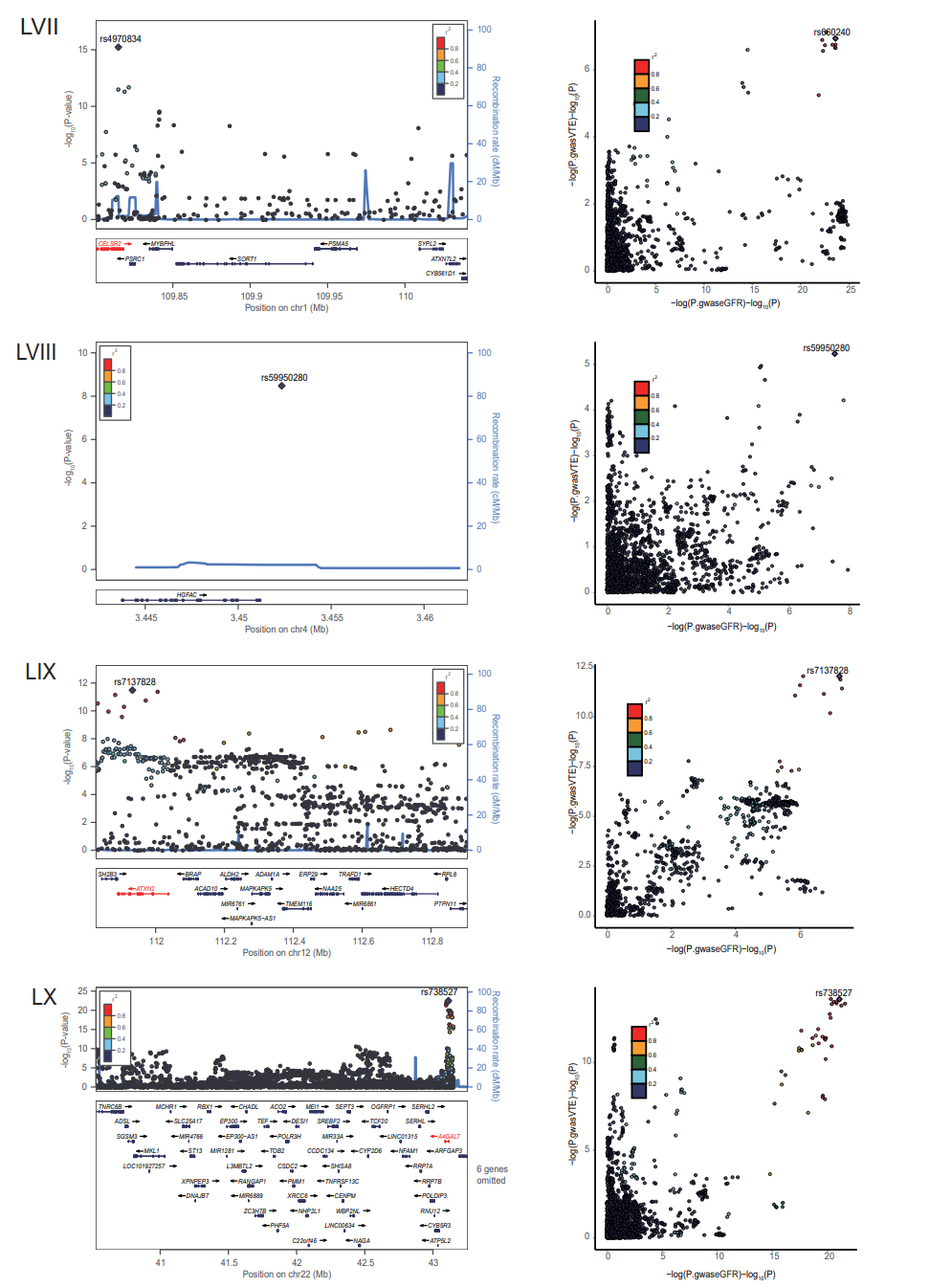

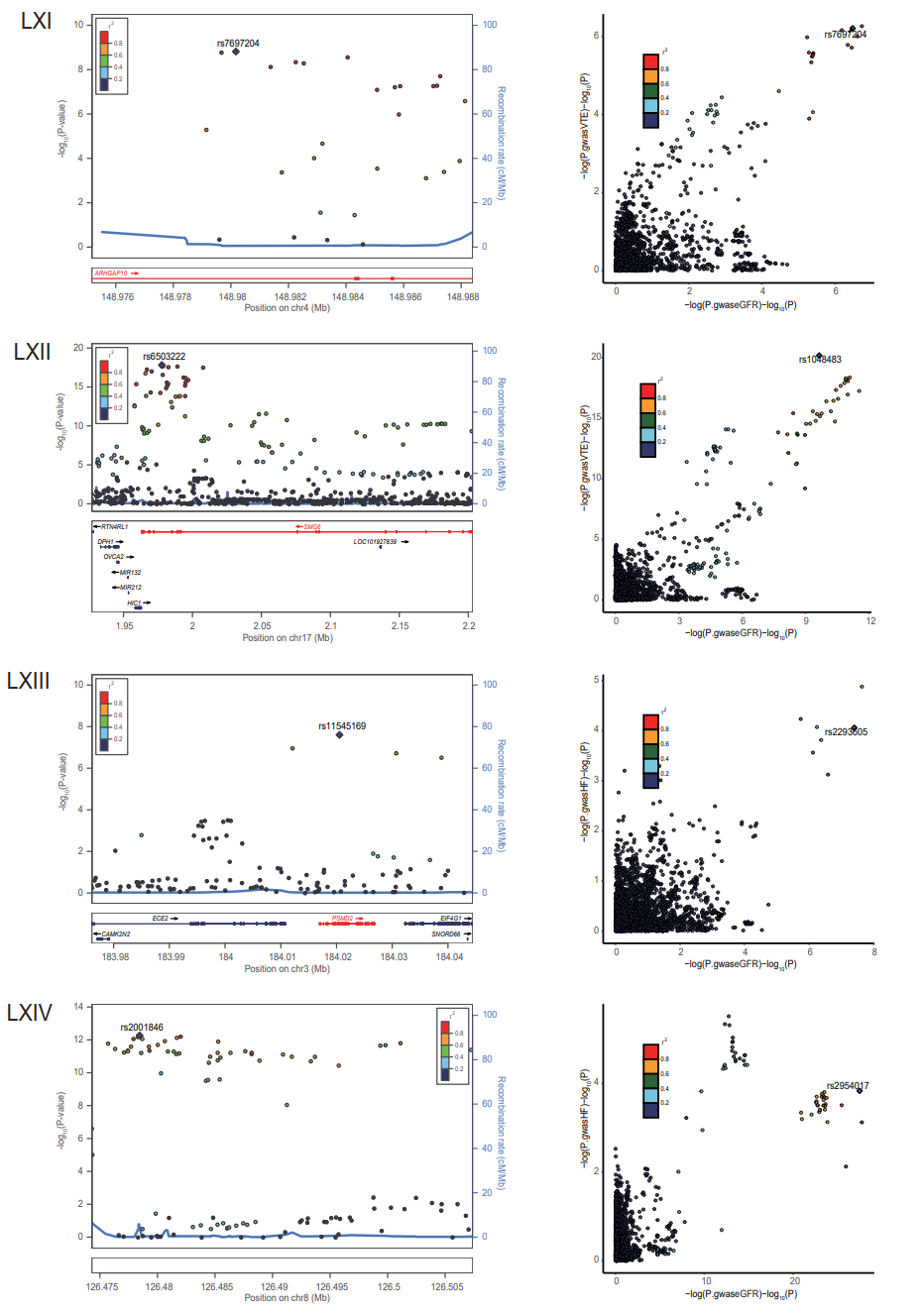

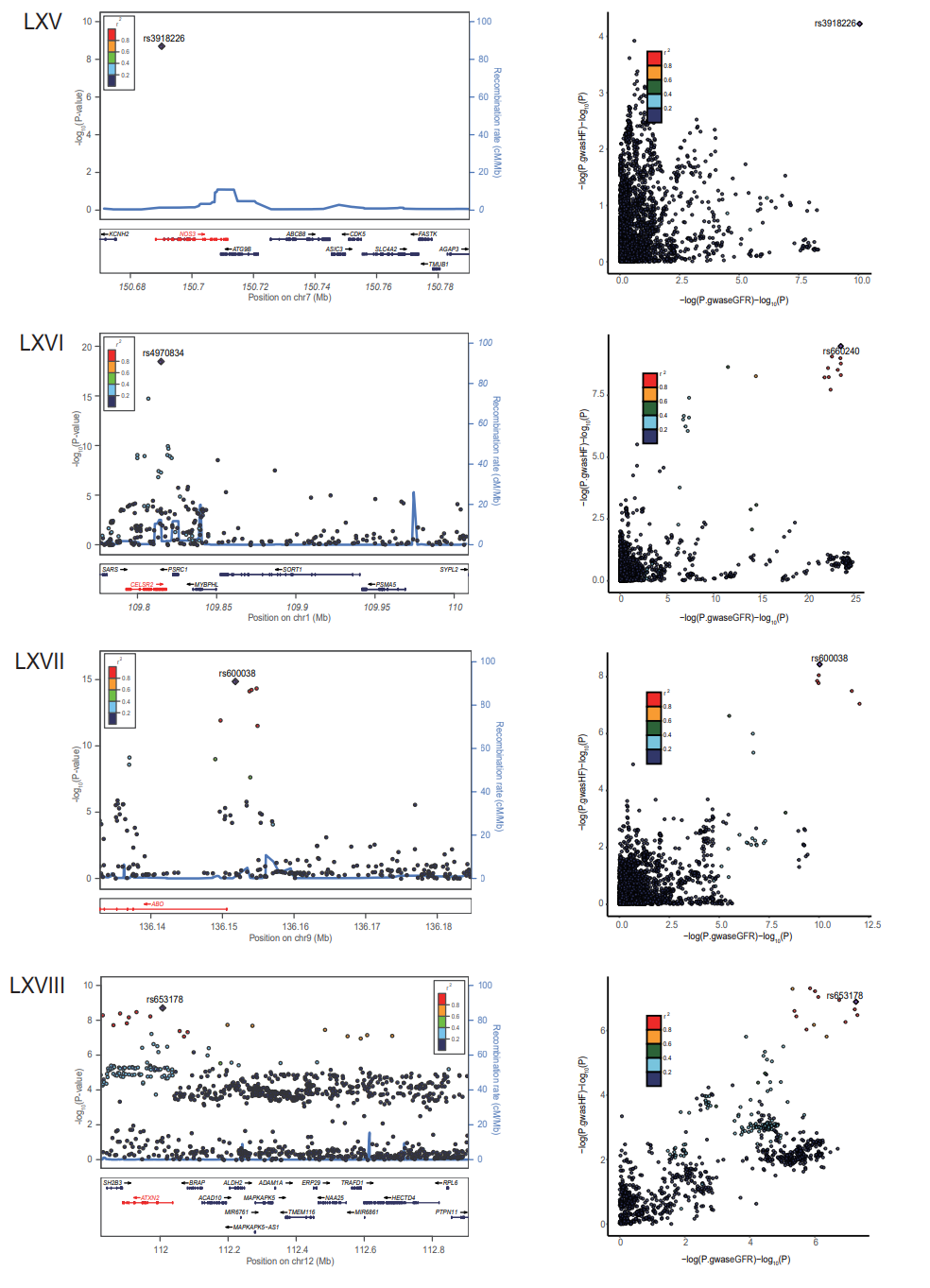

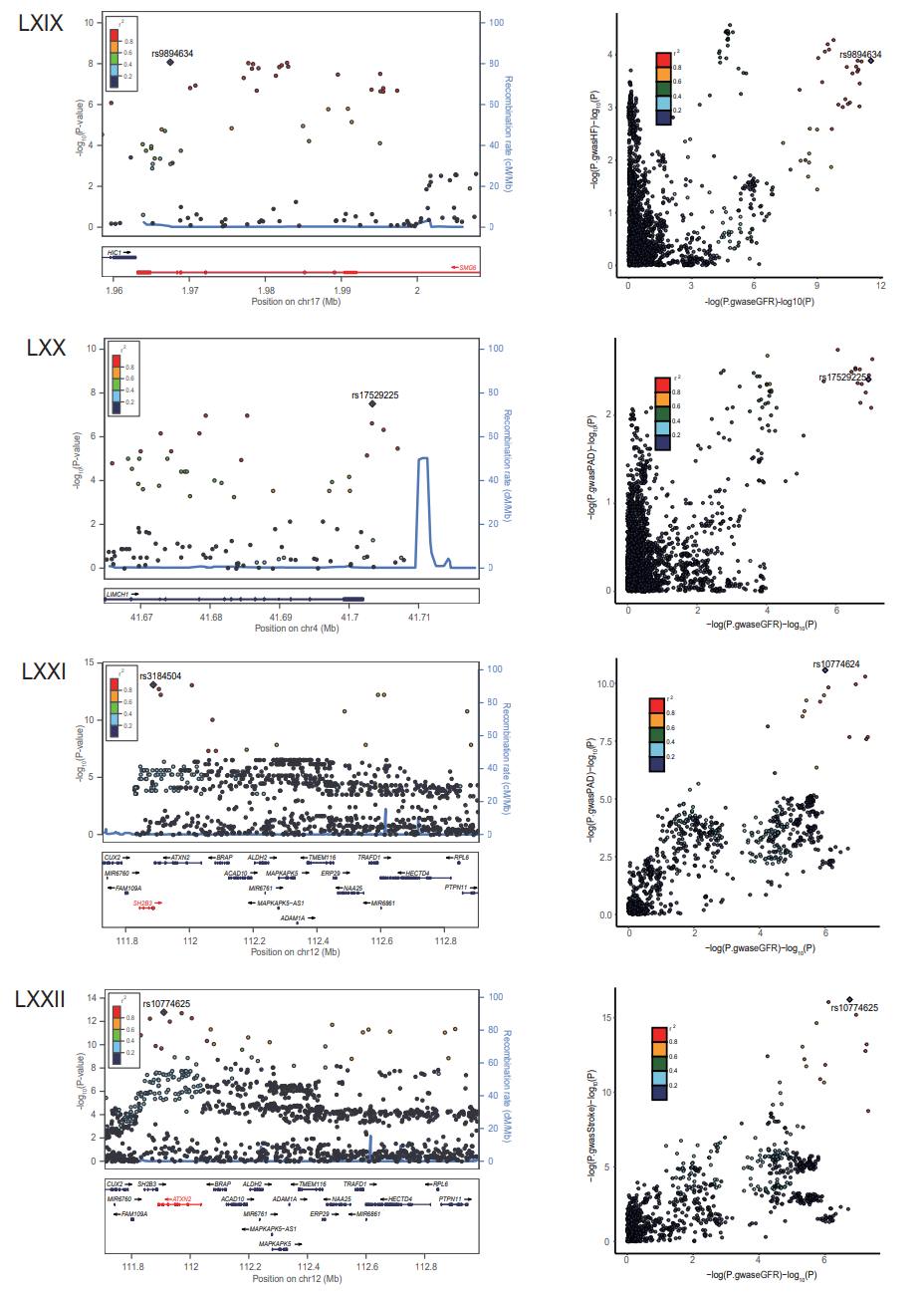

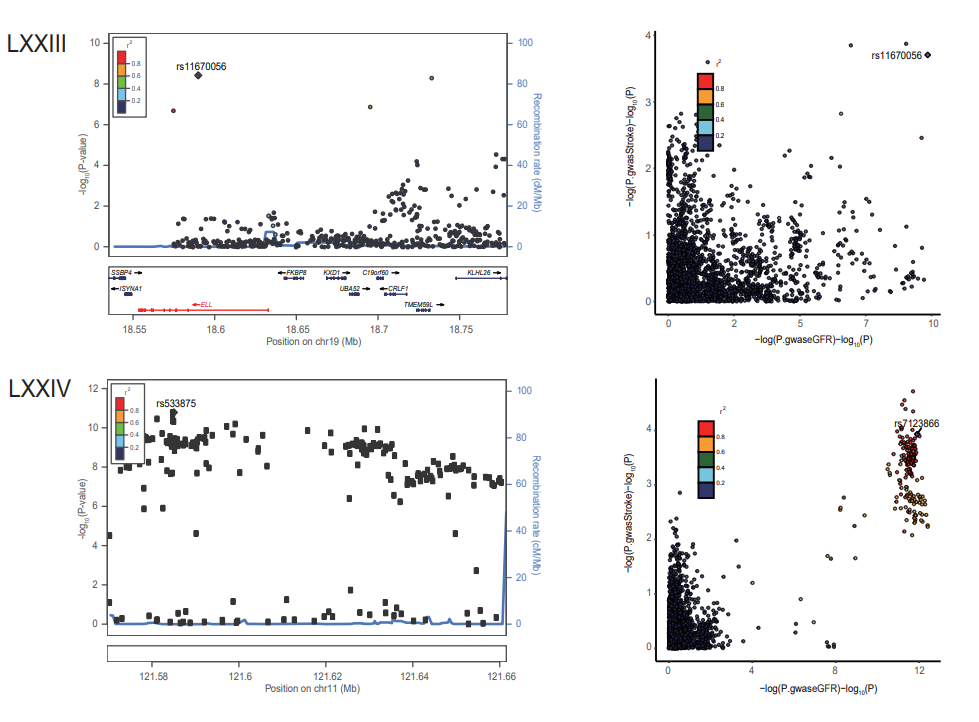

**Supplementary Figure 7: Locus comparison plots of shared causal variants associated with estimated glomerular filtration rate and six major cardiovascular diseases.**

A total of 74 genomic loci were identified to have strong evidence between eGFR and CVDs (PP.H4 > 0.7). The left panel depicts the PLACO results using LocusZoom plots, and the right panel compares the two single-trait GWAS statistics for each variant-trait pair using LocusCompare plots. For the LocusZoom plots, the x-axis shows the genomic position of each variant, and the y-axis shows the -log10 P value from the PLCAO results. A purple diamond represents each locus's top variant with the smallest Ppraco. The color of each variant represents its LD relationship with the top variant. For the LocusCompare plots, each dot represents a variant, and the x-axis shows the -log10 PGWAS from the corresponding GWAS for eGFR, and the y-axis shows the -log10 PGWAS from the corresponding GWAS for CVDs. Purple diamonds also represent candidate-shared causal variants identified by pairwise colocalization analysis. The color of each variant represents its LD relationship with the candidate-shared causal variant. All genomic mapping is based on the reference genome hg19, and LD calculations are based on the European population 1000 genomes project. (I) 1q32.1（rs1044145）in eGFR-AF, (II) 12q22（rs11107114）in eGFR-AF, (III) 1p32.3（rs111517573）in eGFR-AF, (IV) 3p25.2（rs11718898）in eGFR-AF, (V) 8q24.3（rs11784619）in eGFR-AF, (VI) 6p21.31（rs141130394）in eGFR-AF, (VII) 6q16.3（rs17789218）in eGFR-AF, (VIII) 4q21.21（rs2903657）in eGFR-AF, (IX) 7q11.21（rs4433015）in eGFR-AF, (X) 18q23（rs4799053）in eGFR-AF, (XI) 12q24.31（rs4930724）in eGFR-AF, (XII) 9q34.2（rs579459）in eGFR-AF, (XIII) 4q21.1（rs62303262）in eGFR-AF, (XIV) 15q26.3（rs6598541）in eGFR-AF, (XV) 18q21.1（rs9953366）in eGFR-AF, (XVI) 19p13.11（rs10401969）in eGFR-CAD, (XVII) 6p21.2（rs10947786）in eGFR-CAD, (XVIII) 12q13.3（rs11609805）in eGFR-CAD, (XIX) 3p21.31（rs11706370）in eGFR-CAD, (XX) 7q36.1（rs117564322）in eGFR-CAD, (XXI) 12q24.31（rs12301673）in eGFR-CAD, (XXII) 1q25.3（rs12739316）in eGFR-CAD, (XXIII) 17q23.1（rs1292043）in eGFR-CAD, (XXIV) 2q37.1（rs12991088）in eGFR-CAD, (XXV) 12q24.12（rs17696736）in eGFR-CAD, (XXVI) 12q13.13（rs2277383）in eGFR-CAD, (XXVII) 17q23.2（rs28394355）in eGFR-CAD, (XXVIII) 17q24.2（rs2909220）in eGFR-CAD, (XXIX) 8q24.13（rs2954021）in eGFR-CAD, (XXX) 3q25.2（rs357494）in eGFR-CAD, (XXXI) 4q21.21（rs36034102）in eGFR-CAD, (XXXII) 4q22.3（rs3775058）in eGFR-CAD, (XXXIII) 7p21.1（rs4410790）in eGFR-CAD, (XXXIV) 4q22.3（rs4699540）in eGFR-CAD, (XXXV) 1p13.3（rs55882046）in eGFR-CAD, (XXXVI) 2p23.3（rs55952936）in eGFR-CAD, (XXXVII) 4p16.3（rs59950280）in eGFR-CAD, (XXXVIII) 9q34.2（rs651007）in eGFR-CAD, (XXXIX) 5q11.2（rs72750233）in eGFR-, (XL) 6q15（rs72912510）in eGFR-CAD, (XLI) 19p13.11（rs78030362）in eGFR-CAD, (XLII) 1p36.11（rs79598313）in eGFR-CAD, (XLIII) 20q11.22（rs945959）in eGFR-CAD, (XLIV) 17q12（rs9652838）in eGFR-CAD, (XLV) 1p22.2（rs10922478）in eGFR-VTE, (XLVI) 2q34（rs12151812）in eGFR-VTE, (XLVII) 1q44（rs12403095）in eGFR-VTE, (XLVIII) 3p22.2（rs1274961）in eGFR-VTE, (XLIX) 1p36.11（rs12753968）in eGFR-VTE, (L) 22q12.3（rs132648）in eGFR-VTE, (LI) 3p24.2（rs2306623）in eGFR-VTE, (LII) 5q14.3（rs34580448）in eGFR-VTE, (LIII) 19q13.32（rs34783010）in eGFR-VTE, (LIV) 3q25.2（rs357468）in eGFR-VTE, (LV) 19q13.11（rs41523449）in eGFR-VTE, (LVI) 12q24.31（rs4930724）in eGFR-VTE, (LVII) 1p13.3（rs4970834）in eGFR-VTE, (LVIII) 4p16.3（rs59950280）in eGFR-VTE, (LIX) 12q24.12（rs7137828）in eGFR-VTE, (LX) 22q13.2（rs738527）in eGFR-VTE, (LXI) 4q31.23（rs7697204）in eGFR-VTE, (LXII) 17p13.3（rs9901671）in eGFR-VTE, (LXIII) 3q27.1（rs11545169）in eGFR-HF, (LXIV)8q24.13（rs2001846）in eGFR-HF, (LXV) 7q36.1（rs3918226）in eGFR-HF, (LXVI) 1p13.3（rs4970834）in eGFR-HF, (LXVII) 9q34.2（rs600038）in eGFR-HF, (LXVIII) 12q24.12（rs653178）in eGFR-HF, (LXIX) 17p13.3（rs9894634）in eGFR-HF, (LXX) 4p13（rs17529225）in eGFR-PAD, (LXXI) 12q24.12（rs3184504）in eGFR-PAD, (LXXII) 12q24.12（rs10774625）in eGFR-Stroke, (LXXIII) 19p13.11（rs11670056）in eGFR-Stroke, (LXXIV) 11q24.1（rs533875）in eGFR-Stroke. Detailed description is shown in Supplementary Table 7.
